# Adeno-associated virus 2 infection in children with non-A-E hepatitis

**DOI:** 10.1101/2022.07.19.22277425

**Authors:** Antonia Ho, Richard Orton, Rachel Tayler, Patawee Asamaphan, Lily Tong, Katherine Smollett, Chris Davis, Maria Manali, Sarah E. McDonald, Louisa Pollock, Clair Evans, Jim McMenamin, Kirsty Roy, Kimberly Marsh, Titus Divala, Matt Holden, Michael Lockhart, David Yirrell, Sandra Currie, Samantha J. Shepherd, Celia Jackson, Rory Gunson, Alasdair MacLean, Neil McInnes, Richard Battle, Jill Hollenbach, Paul Henderson, Meera Chand, Melissa Shea Hamilton, Diego Estrada-Rivadeneyra, Michael Levin, DIAMONDS consortium, ISARIC4C Investigators, David L Robertson, Ana Filipe, Brian Willett, Judith Breuer, Malcolm G Semple, David Turner, J Kenneth Baillie, Emma C. Thomson

## Abstract

An outbreak of acute hepatitis of unknown aetiology in children was first reported in Scotland in April 2022.^1^ Cases aged <16 years have since been identified in 35 countries.^2^ Here we report a detailed investigation of 9 early cases and 58 control subjects. Using next-generation sequencing and real-time PCR, adeno-associated virus 2 (AAV2), was detected in the plasma of 9/9 and liver of 4/4 patients but in 0/13 sera/plasma of age-matched healthy controls, 0/12 children with adenovirus (HAdV) infection and normal liver function and 0/33 children admitted to hospital with hepatitis of other aetiology. AAV2 typically requires a coinfecting ‘helper’ virus to replicate, usually HAdV or a herpesvirus. HAdV (species C and F) and human herpesvirus 6B (HHV6B) were detected in 6/9 and 3/9 affected cases, including 3/4 and 2/4 liver biopsies, respectively. The class II HLA-DRB1*04:01 allele was identified in 8/9 cases (89%), compared with a background frequency of 15.6% in Scottish blood donors, suggestive of increased susceptibility in affected cases. Acute non-A-E paediatric hepatitis is associated with the presence of AAV2 infection, which could represent a primary pathogen or a useful biomarker of recent HAdV or HHV6B infection. Population and mechanistic studies are required to explore these findings further.

## Main text

### Hepatitis outbreak in Scottish children

In April 2022, a cluster of five young children presented with jaundice^i^ and acute severe hepatitis of unknown aetiology were reported by several hospital sites in Scotland.^1^ In response, Public Health Scotland (PHS) convened a National Incident Management Team (IMT), alongside the UK Health Security Agency (UK HSA) and academic partners (ISARIC4C Investigators) to coordinate further investigations and the public health response. As of 4 July 2022, 36 children aged 10 years and under with acute unexplained non-A to E hepatitis have been confirmed in Scotland, one of whom required liver transplantation (**Figure 1a**).^3^ Elsewhere in the UK, 186 confirmed cases have been reported and 14 children have required liver transplantation.^3^ Globally, the World Health Organisation (WHO) has reported over 1010 probable cases fulfilling their case definition in 35 countries.^2^ Clinical investigations in Scotland excluded all expected causes of acute hepatitis including hepatitis A, B, C and E viruses, HSV, CMV and EBV. Adenovirus (HAdV) was identified by PCR in 53.1% (17/32) of tested cases. Recent reports from the UK and the USA have also identified HAdV to be present in the majority of cases in two cohorts of children^4–6^. A spike of HAdV infections in Scotland predominantly affecting children <5 years also directly preceded the outbreak of unexplained hepatitis (**Figure 1b**). SARS-CoV-2 PCR was positive in 16.7% (6/36 tested cases).^3^ A rise in COVID-19 cases in the community preceded the spike in non-A-E hepatitis by around two years (**Figure 1c**). Human herpesvirus 6 (HHV6A and HHV6B) infections were not detected at higher levels during 2021 or 2022 (**Figure 1d**).

**Figure 1:**
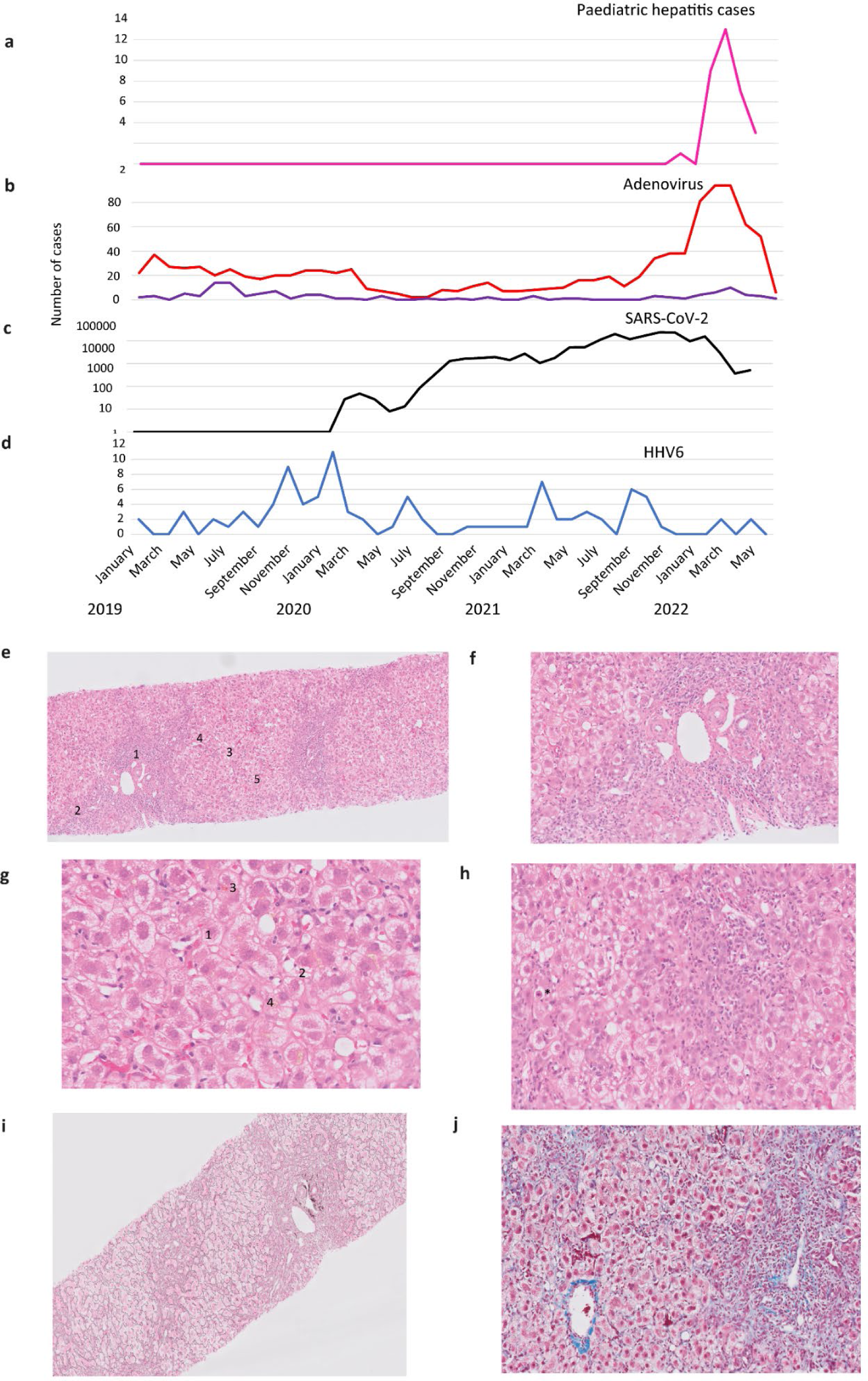
Epidemiology and histology of liver biopsies in Scottish children with AAV2 and HAdV-associated hepatitis. (a) Emergence of acute non-A to E hepatitis in children Cases of non-A-E hepatitis in children 10 years and under in Scotland, 11 January to 20 June 2022 (pink).^3^ Cases of HAdV (children <5 years: red, children 5-10 years: purple) (b), SARS-CoV-2 (black) (c) and HHV6 (blue) (d) in children aged 10 years and under in Scotland January 2019 to June 2022. The histology of Case 3 (CVR3) is shown in panels (e-j). A low-power liver biopsy image (e) showing expansion of the portal tracts by inflammatory cells (1), interface hepatitis (2), feathering (ballooning) change of hepatocytes (3) and apoptotic hepatocytes (4). Mild steatosis is present in some zone 2 hepatocytes (5). A medium power view of the same biopsy (f) illustrates an affected portal tract showing a mixed infiltrate of inflammatory cells (lymphocytes, plasma cells, eosinophils and neutrophils). Vessels and bile ducts are preserved. A high-power picture (g) of the liver parenchyma showing feathering (ballooning) change in the hepatocytes (1), focal mild steatosis (2), focal bile canaliculi plugging (3) and lipofuscin pigment (4). An apoptotic hepatocyte (h) is highlighted with an asterisk *. A reticulin stain of the same biopsy (i) highlights an increased density of fibres in areas of inflammation and some change of the zone 2 parenchyma. A Masson trichrome stain showing collagen in blue (j) within portal tracts and mild staining extending around some hepatocytes.

### Research investigation

To investigate the aetiology of the acute hepatitis cases, we recruited nine of the earliest affected children who presented to hospital between 14 March and 4 April 2022 into the International Severe Acute Respiratory and Emerging Infections Consortium (ISARIC) WHO Clinical Characterisation Protocol UK (CCP-UK) [ISRCTN 66726260]^7^. Ethical approval was given by the South Central–Oxford C Research Ethics Committee in England (13/SC/0149), the Scotland A Research Ethics Committee (20/SS/0028), and the WHO Ethics Review Committee (RPC571 and RPC572). Control samples were obtained from the Diagnosis and Management of Febrile Illness using RNA Personalised Molecular Signature Diagnosis study cohort (DIAMONDS; https://www.diamonds2020.eu). This study recruited children presenting with suspected infection or inflammation. Patients were recruited with the informed written consent of parents or guardians. The DIAMONDS study was approved by the London – Dulwich Research Ethics Committee (20/HRA/1714).

Control subjects were restricted to children recruited in the UK between January 2020 and April 2022. Three comparison groups were identified: Group 1, sera from 13 age-matched healthy children (10 male, 3 female; age range 3-5 years); Group 2, 12 children (8 male, 4 female; age range 1-4 years) with PCR-confirmed HAdV infection and normal transaminases (n=12); Group 3, 33 children (18 male, 15 female; age range 2-16 years) with raised transaminases who were HAdV PCR negative. Within Group 3, 15 children required critical care for ventilatory or cardiovascular support. There was no significant difference in age between cases and healthy controls, but some control samples were sampled earlier than case samples (January 2020-April 2022 versus March-April 2022). Children with HAdV infection with normal liver function were younger (median age 1.6 years; interquartile range (IQR) 1.1-3.3 years, p<0.001), and children with deranged liver function and no adenoviral infection were older (median age 9.2 years; IQR 6.7-13.6 years, p=0.001), compared to cases. (**Supplementary Table 1**).

### Clinical presentation

The median age of case patients was 3.9 years (IQR: 3.4 to 5.1 years). Four of the nine children were male, and all were of white ethnicity. All were admitted to hospital and met the PHS case definition for confirmed acute hepatitis, with a serum transaminase >5As the epidemiology was in keeping 00 IU/L (AST or ALT) without any known cause, aged 10 years or under and presenting after 1st January 2022. The majority of the children reported a preceding subacute history of abdominal pain, diarrhoea and vomiting, occurring between 1 to 11 weeks prior to acute hepatitis. None of the cases were immunocompromised and none had received COVID-19 vaccination. All routine blood tests for viral hepatitis, including hepatitis A, B, C, E, Epstein-Barr virus (EBV), cytomegalovirus (CMV), human herpes virus (HHV) 6/7 and herpes simplex virus (HSV) were negative (**Supplementary Table 2**). One case had weakly positive antinuclear antibodies (ANA; titre 1:80), but otherwise autoimmune markers, [immunoglobulins, antinuclear antibodies (ANA), anti-mitochondrial antibodies (AMA), anti-smooth muscle antibodies (ASMA), anti-liver-kidney microsomal (anti-LKM) antibody and tissue transglutaminase antibody (TTG)] were negative. Four of the children had liver biopsies, which all showed evidence of lobular hepatitis with periportal and interface inflammation, bile duct proliferation and ballooning of hepatocytes, but no evidence of confluent fibrosis (**Figures 1e-j**). Two cases required transfer to a specialist liver unit due to significant synthetic liver dysfunction. Both improved with steroid therapy and neither required liver transplantation. There were no deaths and the median duration of hospital stay was 10 days (range 5-32 days).

### Genome sequencing

As the epidemiology was in keeping with the emergence of an infectious pathogen, we undertook metagenomic and target enrichment sequencing and real-time PCR on all available clinical samples of cases, including plasma (n=9), liver biopsies (n=4), throat swabs (n=6), faecal samples (n=7) and a rectal swab (n=1), obtained between 7 to 80 days from initial symptom onset (**Figures 2a-d**, **Supplementary Tables 3-5**). Metagenomic sequencing was carried out using protocols designed to identify both RNA and DNA viruses. Semi-agnostic target enrichment sequencing was also performed using VirCapSeq-VERT Capture probes that target the genomes of 207 viral taxa known to infect vertebrates (**Supplementary Information**). The most frequently detected viral genomes in affected patient plasma were adeno-associated virus 2 (AAV2) in 9/9 patients (range 152-24,968 sequence reads per million) and at a lower read count, HAdV-F41 or HAdV-C (types 1, 2, 5 and 6 could not be distinguished reliably due to low read counts) in 6/9 patients (range 0-1.66 reads per million). HHV6B was detected at low level in the liver biopsy samples of 2/4 patients by PCR and 3/4 plasma samples by NGS but was not confirmed by PCR in plasma in 9/9 patients (**Figures 2e, 3a**).

**Figure 2.**
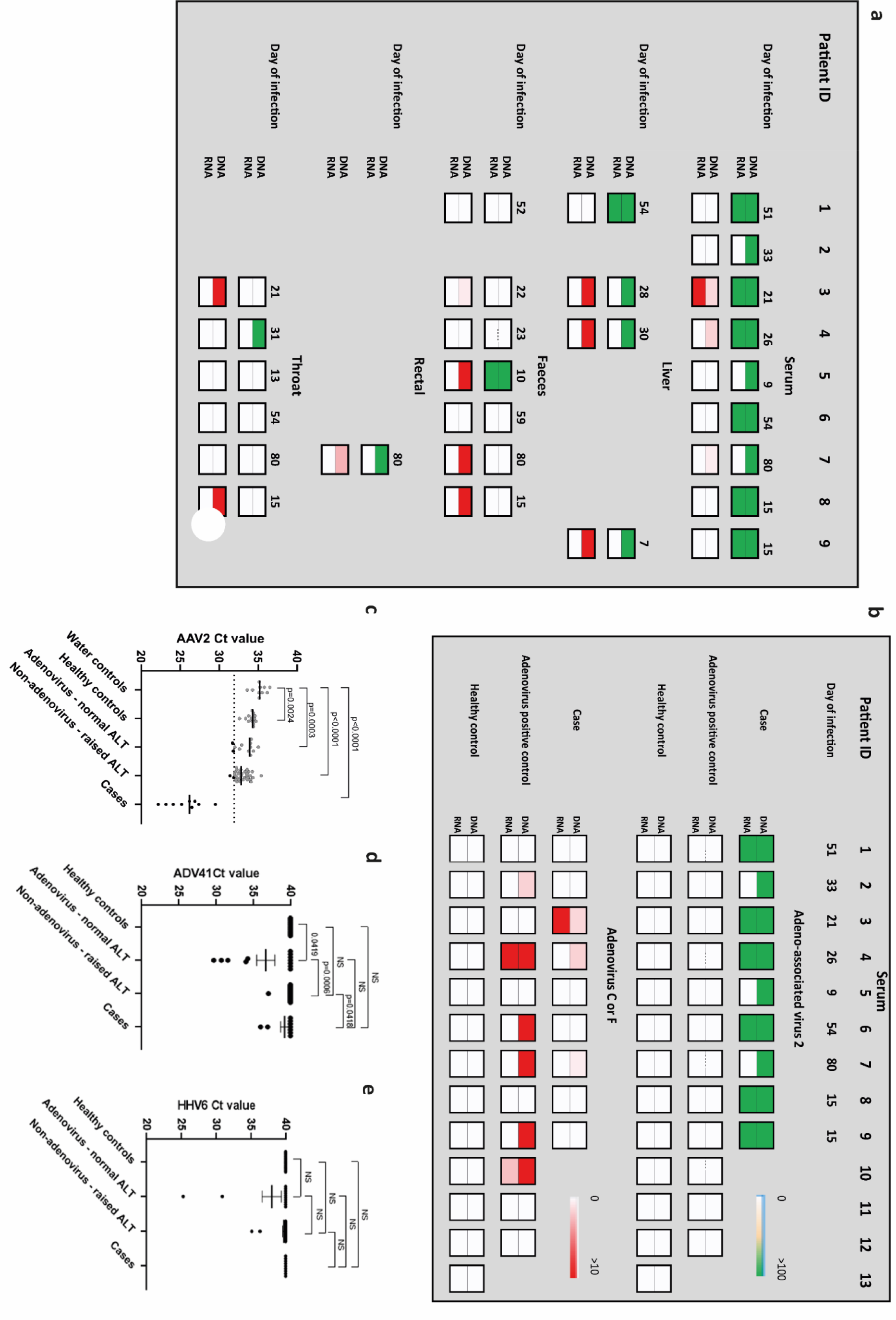
(a) Heatmap of HAdV and AAV-2 reads detected in hepatitis cases by target enrichment sequencing. Samples obtained for routine clinical investigation (plasma, liver, faeces, rectal and throat swab) were retrospectively sequenced following DNA or RNA extraction. AAV2 read counts are shown from 0->100 reads/million in green (upper row) and HAdV read counts are shown from 0->10 reads/million in red (lower row). **(b) Heatmap of viral reads from plasma in cases and plasma/sera from controls.** Plasma samples from cases, and plasma or sera samples from children with HAdV infection and age-matched healthy controls were sequenced following DNA or RNA extraction. AAV2 read counts are shown from 0->100 reads/million in green and HAdV read counts are shown from 0->10 reads/million in red. The number of days between initial symptom onset and sample are indicated. **(c) AAV2 real-time PCR** in cases versus controls. The estimated detection limit of the assay (Ct=31.9) is shown (dotted line) Error bars represent median. Statistical analysis was performed using Mann-Whitney test. **(d) HAdV41 real-time PCR** in cases versus controls. **(e) HHV6 real-time PCR** in cases versus controls.

**Figure 3.**
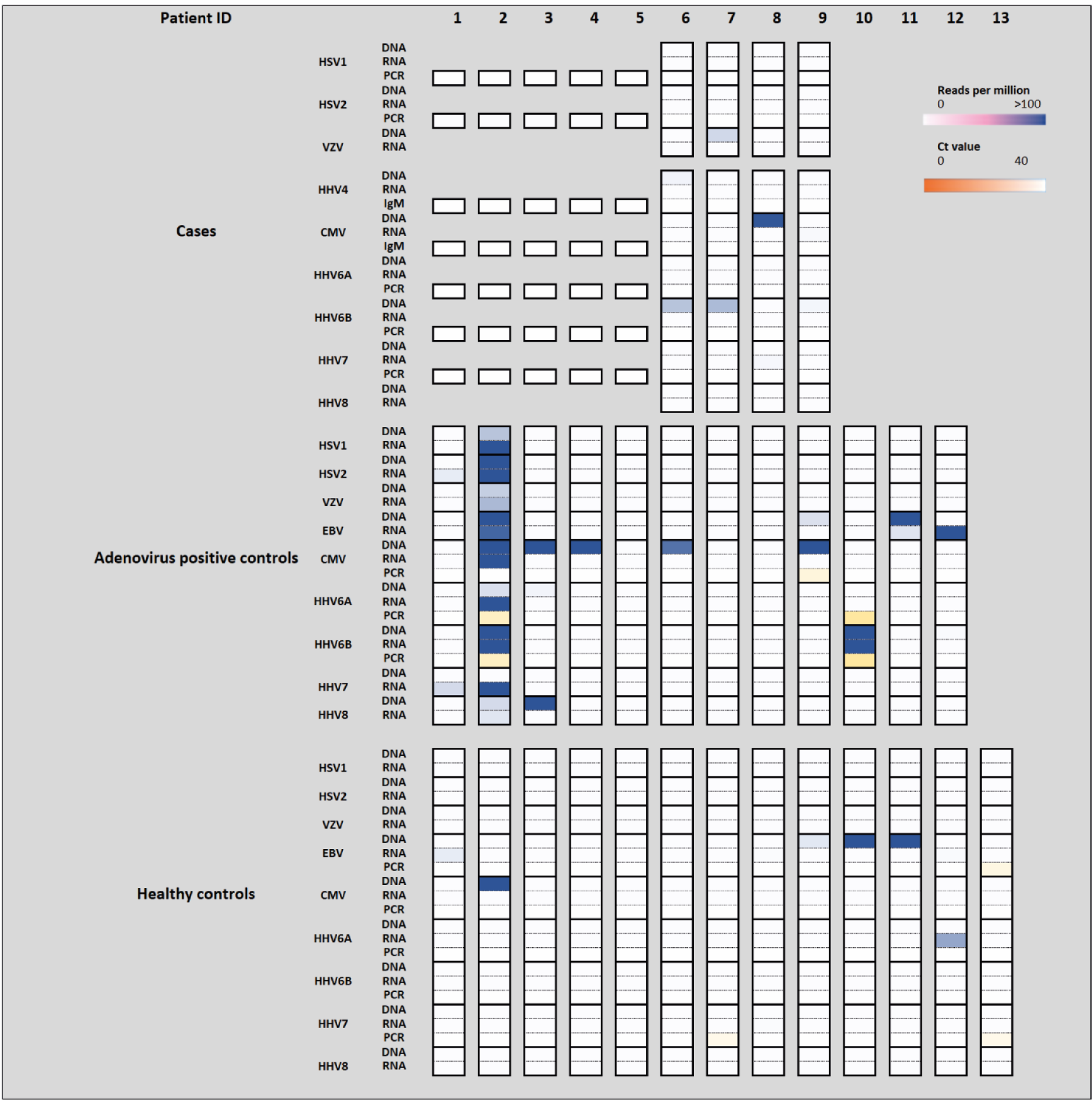
Herpesviruses detected in cases and controls. **(a)** Results are shown from target enrichment sequencing, PCR and IgM serology. Sequencing read counts are shown in blue as a heatmap from 0-100 reads per million. Ct values are highlighted in orange from 0-40. As the plasma samples from patients 1-5 had a Murid herpesvirus as a spike in control, the herpesvirus content of these samples was not analysed.

AAV2 was detected in 9/9 serum samples and 4/4 liver biopsies, 1/7 faecal, 1/1 rectal and 1/6 throat swab samples. Near full genomes of AAV2 were detected in all 9 patients and contained two large open reading frames corresponding to rep and cap, flanked by ITR regions. Seven distinct lineages of AAV2 were noted (**Figure 4a**) within a cluster containing 4 AAV2 genomes detected in France between 2004 and 2015. Two of three identical sequences were known to have come from a single household and the third occurred around the same time but was not known to be otherwise linked. Several mutations within the VP1-3 genes were noted to be over-represented in the sequences derived from patients with hepatitis when compared with reference sequences (**Figure 4c**). Notably, 9 of the capsid gene mutations over-represented in these cases (V151A, R447K, T450A, Q457M, S492A, E499D, F533Y, R585S and R588T) are associated with a sub-lineage of AAV2 (AAVv66) that has an altered phenotype, including substantial evasion of neutralising antibodies directed against wild-type AAV2, enhanced production yields, reduced heparin binding and increased virion stability. Intrathecal injection of AAVv66 in mice resulted in widespread CNS transduction compared with more localised spread in mice injected with wildtype AAV2.^8^

**Figure 4.**
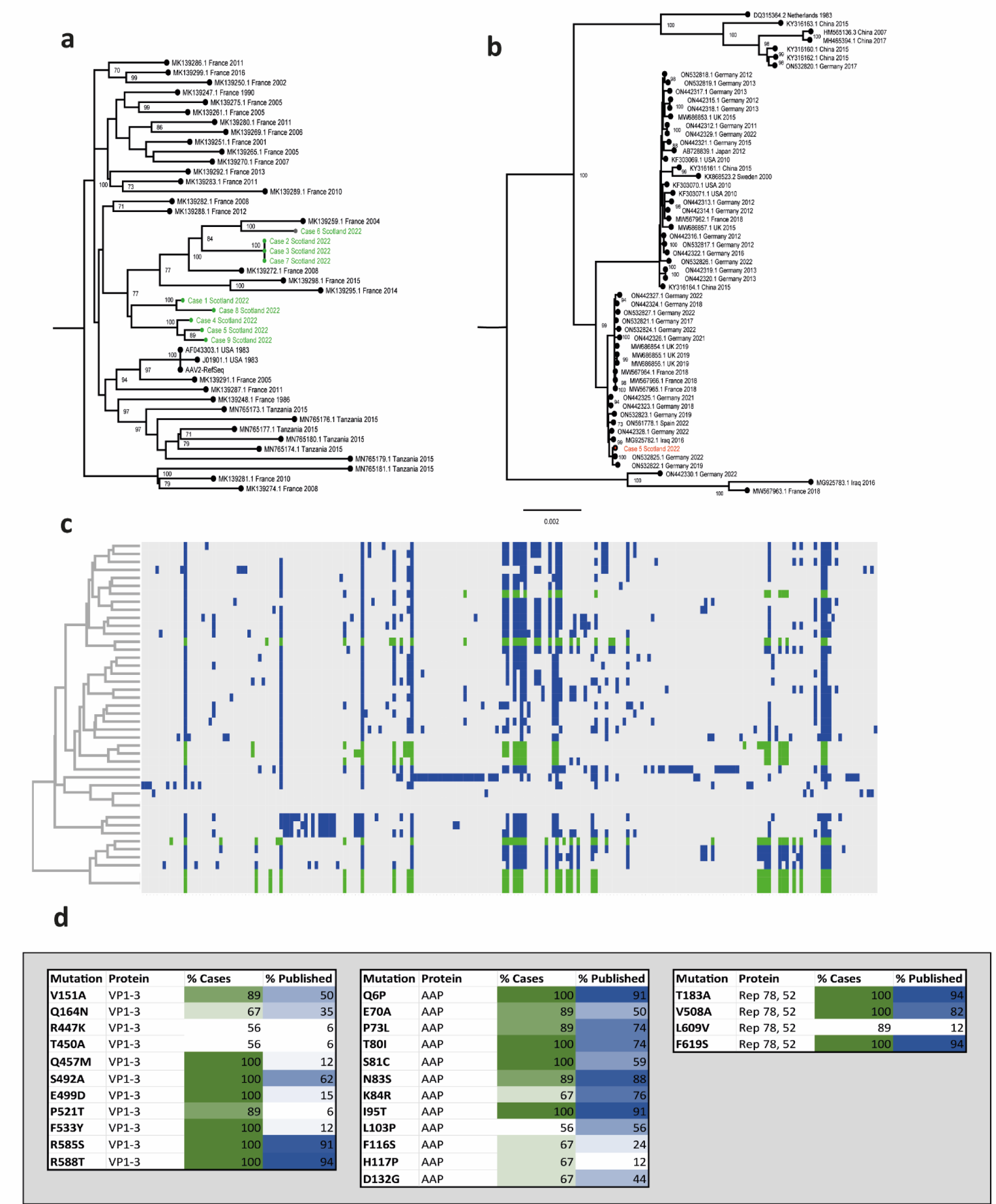
(a) Phylogeny of AAV2 genomes from cases 1-9. The nine AAV2 genome sequences generated from the plasma samples via target enrichment (highlighted in red) were aligned with a range of the closest AAV sequences from La Bella *et al*.^25^ and Cordey *et al*.^26^ IQ-TREE 2 was used to find the optimal model (TIM+F+R3) and create a maximum likelihood (ML) phylogenetic tree with 1000 bootstrap replicates (displayed on the branches). The tree is midpoint rooted. AAV2 reference sequences are denoted by accession number, country and year of sampling **(b) Phylogeny of HAdV41 genome from case 5** A near-full HAdV41 genome sequence from the faecal sample of patient 5 (highlighted in red) was combined with complete genomes of HAdV41 from GenBank. After alignment with MAFFT, IQ-TREE 2 was used to determine the optimal evolutionary model (K2P+R2) and to construct a ML phylogenetic tree with 1000 bootstrap replicates. The tree is midpoint rooted. HAdV41 reference sequences are denoted by accession number, country and year of sampling. (c) **Key mutations and hierarchical clustering of AAV2 genomes** Mutations in published AAV2 sequences are highlighted in (blue) and case sequences (green) **(d) Mutations over-represented in cases versus controls** Mutations in VP1-3, Rep78 and 52 and AAP are highlighted by % representation in case sequences (green) and published sequences (blue)

HAdV was detected in 3/9 plasma samples, 3/4 liver biopsies, 2/6 throat swabs, 4/7 faecal samples and 1/1 rectal swab (**Figure 2b; Supplementary Table 3**). The lower detection of HAdV by PCR compared to enrichment sequencing likely reflects slightly lower sensitivity of the PCR assay. However low numbers of positive samples using both assays may reflect the fact that plasma is a suboptimal sample type for HAdV detection (whole blood samples were unavailable). A full genome of HAdV-F41 was retrieved from a faecal sample (accession number pending) and was found to be closest to two genomes reported from Germany in 2019 and 2022 (**Figure 2b**). As described above, HHV6B was detected at low read number (0.41- 3.89 reads per million) in 3 out of 4 plasma samples (**Figure 3a**, **Supplementary Table 4**). The remaining clinical samples were excluded from analysis for HHV by sequencing because murine CMV had been added as an extraction control during routine clinical investigation. All 9 plasma samples tested negative by PCR for HHV6, HSV, CMV and EBV. Two liver biopsy specimens tested positive for HHV6 (Ct 33 and 36). Contigs matching to other known human pathogens, including human coronavirus NL63, rhinovirus C, enterovirus B, human parainfluenza virus 2 and 3, norovirus, as well as other beta- and gammaherpesviruses were also detected although not consistently across cases (**Supplementary Table 2**). These were detected by both sequencing and PCR.

To investigate the presence of AAV2 and the candidate helper viruses HAdV and HHV6B in case samples, we undertook a series of case-control studies using plasma by sequencing and by real-time PCR (**Figures 2b-e, 3a**). Higher read counts of HAdV (p=0.0545), AAV2 (p<0.001) and HHV6B (p=0.0059) were found in cases versus healthy controls (**Supplementary Figures 1a,1b**). Metagenomic and target enrichment sequencing from the 13 age-matched healthy control samples (group 1) revealed no evidence of AAV2, HAdV or HHV6B in plasma, although low read counts of EBV, CMV and HHV6A were detected (**Figure 3a, Supplementary Figure 1c-j**)). 12 HAdV-infected controls (10 diagnosed by nasopharyngeal aspirate, 1 by nose swab and 1 by stool) with normal liver function (group 2) were found to have no evidence of AAV2. HAdV reads were also detected in 6/12 plasma/serum samples, and 9/12 of this group had a herpesvirus detected, including 2/12 with detectable numbers of HHV6B reads (1050-5062 reads per million), confirmed by PCR (**Figure 2a; Supplementary Table 6; Supplementary Figure 1**). In keeping with the possibility of reactivation of HHV6B, 6/12 of this control group had been admitted to hospital with a critical illness.

### Real-time PCR

To quantify the level of AAV2 and to further investigate the presence of the helper viruses HAdV and HHV6B, we carried out real-time PCR in case plasma and control sera/plasma. We also included a third control group of patients with hepatitis of alternative aetiology, half of whom had been admitted to critical care (**Figures 2c-e**). Significance differences between groups for Ct values and viral loads/ml of plasma were calculated using the Mann-Whitney test. AAV2 PCR of plasma from 9/9 cases was positive with a median estimated copy number of 8,058,879 copies/ml (**Figure 2c; Supplementary Figure 2**). 2/9 case plasma were positive for HAdV-F41 by PCR (**Figure 2d**), and all tested negative by PCR for HHV6 DNA (**Figure 2e**).

### SARS-CoV-2 infection

Routine clinical investigation detected SARS-CoV-2 nucleic acid in nasopharyngeal samples from 2/9 children, one of whom was also seropositive. The other developed infection after the onset of hepatitis. SARS-CoV-2 was not detected by PCR and sequencing in any clinical samples, including liver samples, in cases or controls. Nevertheless, to investigate the possibility that unexplained hepatitis in children might relate to a preceding SARS-CoV-2 infection or other seasonal coronaviruses, we carried out serology. IgG antibody titres were measured quantitatively against SARS-CoV-2 trimeric spike (S) protein, N-terminal domain (NTD), receptor binding domain (RBD) and nucleocapsid (N), and the human seasonal coronaviruses (HCoVs) 229E, OC43, NL63 and HKU1. Electrochemiluminescence assay (MSD-ECL) for coronavirus-specific IgG revealed high levels of prior exposure to seasonal coronaviruses; NL63 (9/9) and OC43 (7/9) (**Supplementary Figure 3a**). In comparison, plasma from 5/9 children displayed reactivity with HKU1 and 3/9 with 229E. Plasma from three children reacted with SARS-CoV-2 N, a fourth was considered weak positive in comparison with the reference range of known negative individuals (**Supplementary Figure 3b**). This child was one of 3/4 of the N-positive children displaying reactivity with S, NTD and RBD, confirming likely SARS-CoV-2 exposure. Plasma from the fourth child reacted with N only, while plasma from an additional two children reacted with S, NTD and RBD only (no N reactivity) in keeping with exposure to natural infection in the absence of vaccination. In summary, 6/9 (67%) of the children displayed evidence of prior exposure to SARS-CoV-2; this level was comparable to SARS-CoV-2 seroprevalence in children aged 5-11 years in Scotland between 14^th^ March and 4^th^ April 2022 (time period when clinical samples of cases were obtained), reported as between 59% (95% CI 50.6-71.2) and 67% (56.2-77.6%).^9^

### HLA typing

To investigate the possibility that some children might be more susceptible to non-A-E hepatitis, cases and local controls (all of white ethnicity) were genotyped using high resolution typing for all HLA loci (HLA-A, B, C, DRB1, DRB3/4/5, DQA1, DQB1, DPA1 and DPB1) using the AllType™ FASTplex™ NGS Assay (One Lambda). We analysed HLA allele positivity in the nine cases plus an additional 11 ISARIC CCP-UK study patients (n=20 in total) compared to 64 Scottish platelet apheresis donor controls. Associations were observed for the following alleles; DRB1*04:01 (OR 11.8 (95% CI 4.43-32.13), p = 1.19 x 10^-9^), DQA1*03:03 (OR 10.64 (4.07-28.18), p = 3.8 x 10^-9^) and DQB1*03:01 (OR 6.16 (2.49-15.21), p = 3.75 x 10^-6^). The strength of associations and the known linkage disequilibrium between HLA-DRB1*04:01 and associated alleles at other HLA loci indicates that DRB1*04:01 is the primary HLA associated allele. In patients, the HLA-DRB1*04:01 allele frequency was 0.50 and in local controls was 0.078. As a further reference, in 599,410 British/Irish North-West European (BINWE) individuals on the Anthony Nolan register, the frequency of DRB1*04:01 is 0.114.^10^ Of the nine patients in which metagenomic and target enrichment sequencing was undertaken, 8/9 carried the HLA-DRB1*04:01 allele and one carried the HLA-DR4 allele, DRB1*04:07.

### Conclusions/final statements

AAV2 detected independently by sequencing and by real-time PCR was present at significantly higher frequency in cases of paediatric non-A-E hepatitis than controls. That this occurs in the context of an increased frequency of the DRB1*04:01 allele when compared to a local blood donor control population requires further investigation, but points to a T helper cell-mediated pathological response triggered by exposure to HAdV and/or AAV2 infection.

AAV2 is a small non-enveloped single-stranded DNA virus of 4,675 nucleotides belonging to the species adeno-associated dependoparvovirus A (genus Dependoparvovirus, family Parvoviridae)^11^, first described in 1965 and occurs in up to 80% of the adult population, seroconversion occurring in early childhood following respiratory infection^12^. In one prospective study in the USA, the earliest seroconversion to AAV2 infection occurred in a 9- month-old child and the seroprevalence increased from 24.2% to 38.7% in 3 and 5-year-old children, respectively^13^. This age range coincides with that of the cases in this study, raising the possibility of illness relating to primary infection rather than reactivation. Dependoparvoviruses rely on coinfection with a helper virus for replication, most commonly HAdV or a herpesvirus. The majority of clinical samples taken at presentation with hepatitis were obtained more than 20 days after initial symptom onset, which could explain the absence of a helper virus in some samples, and low viral loads (high Ct value) in positive samples. We detected two candidate helper viruses at low level in the cases; HAdV and HHV6B in 6/9 and 3/9 cases respectively (HHV6 was detected only by NGS at very low read counts and was not confirmed by PCR in any of the cases). HHV6B was also present in two control groups that included children with severe HAdV infection and children with hepatitis of alternative aetiology. As HHV-6 can establish latency and can integrate its genome into the human chromosome, it may reactivate following concomitant illness (or immunosuppression) and may represent either an opportunistic bystander or a pathogen.

We hypothesise that AAV2 may be directly implicated in the pathology of non-A-E hepatitis in children, following transmission as a co-infection with HAdV (or HHV6, although this is much less likely) or as a result of reactivation following HAdV or HHV6 infection. Class II HLA type may contribute to disease susceptibility. In keeping with an immune-mediated aetiology directed against AAV2-infected hepatocytes, hepatitis associated with the emergence of a CD8+ cell-mediated response directed against the AAV2 viral capsid (VP1) was reported in early trials of AAV2 when used as a vector for gene therapy.^14–16^ An alternative explanation is that AAV2 may be a useful biomarker of infection with HAdV (that may be the primary pathogen) that persists for a shorter time than AAV2. Most of our cases developed gastroenteritis symptoms (vomiting and diarrhoea) lasting several days, several weeks before the onset of jaundice. The opportunity to detect virus by sequencing was therefore likely reduced, as these samples were collected at a later stage. Further, whole blood samples would have been likely to increase the sensitivity of detection, but only plasma samples were available. We consider this alternative hypothesis to be less likely because we did not detect AAV2 in a control group of children with HAdV infection who had normal liver function.

HAdV41 is a common cause of diarrhoea in young children^17^ and adenovirus-associated hepatitis has been described, particularly among immunocompromised individuals.^18^ However, it has not previously been associated routinely with severe hepatitis. In the recent outbreaks of unexplained hepatitis in children, it has been inconsistently reported.^4–6, 19–21^ HHV6B is a human herpesvirus that is associated with the childhood illness roseola infantum (sixth disease) and a self-limiting otherwise non-specific febrile illness.^22^ Seroconversion usually occurs by 2 years of age and hepatitis occasionally occurs in immunocompetent individuals.

We investigated the possibility that unexplained cases of hepatitis may be linked to prior COVID-19 infection. Direct SARS-CoV-2 liver injury seems unlikely, since few of our cases (2 of 9) were SARS-CoV-2 PCR-positive on admission, and we did not identify SARS-CoV-2 by PCR or sequencing in any of the clinical samples from cases, including liver biopsies. Further, the SARS-CoV-2 antibody positivity in hepatitis cases was within community positivity rates at that time.^3^ This is in keeping with a case-control analysis by UKHSA reporting no difference in SARS-CoV-2 PCR positivity between hepatitis cases and children presenting to emergency departments between January and June 2022.^3^ Nevertheless, we cannot fully exclude a post-COVID-19 immune-mediated phenomenon in susceptible children.

There are several limitations to this study. Firstly, the presence of AAV2 in cases but not controls may have arisen due to seasonal variation in AAV2 transmission, as some controls were sampled earlier than cases. Secondly, the presence of AAV2 in cases is an association and may not represent direct aetiology; rather the AAV2 may be a useful biomarker of recent HAdV (or less likely HHV6B) infection. We do not consider it likely that AAV2 simply represents a marker of liver damage because it was not present in cases of severe hepatitis of alternative aetiology. Thirdly, while the strong association of the HLA-DRB1*04:01 allele, known to be associated with autoimmune hepatitis^23^ and extra-articular manifestations of rheumatoid arthritis,^24^ may represent a disease susceptibility gene, local population structure could result in an over-estimate of significance. Nonetheless it is plausible that an HAdV infection, along with a coinfecting or reactivated AAV2 infection has resulted, for a proportion of children who are more susceptible (due to the class II HLA allele HLA-DRB1*04:01), in a more severe outcome than might normally be expected for these commonly circulating viruses. The recent clustering of cases may have arisen in part because of changes in exposure pattern to HAdV, AAV2 and HHV6B as a result of the COVID-19 pandemic. Faecal HAdV usually follows seasonal trends with maximal rates of detection in younger age groups. However, the circulation of respiratory viruses was interrupted in 2020 by the implementation of non-pharmaceutical interventions, including physical distancing and travel restrictions, instituted to mitigate SARS-CoV-2 transmission. These measures may have created a pool of susceptible younger children, resulting in much higher rates of HAdV and potentially AAV2 circulation in this population of naïve children when the COVID-19 restrictions were relaxed.

Larger case-control studies are urgently needed to investigate the role of AAV2, HAdV, HHV6B and HLA type in the aetiology of unexplained non-A-E paediatric hepatitis. Retrospective testing of samples from cases of unexplained hepatitis in children is also indicated. Ongoing work to measure IgM and IgG responses against AAV2, HAdV and HHV6B may help to delineate whether the AAV2 has been acquired as a primary co-infection or reactivation.

## Methods

### ISARIC CCP-UK recruitment & DIAMONDS/ study

Ethical approval for the ISARIC CCP-UK study was given by the South Central–Oxford C Research Ethics Committee in England (13/SC/0149), the Scotland A Research Ethics Committee (20/SS/0028), and the WHO Ethics Review Committee (RPC571 and RPC572). Children aged <16 years were recruited by written informed consent (parent or guardian) from the ISARIC WHO CCP-UK cohort admitted to hospital with elevated transaminases (defined as ALT >400 iU/L and/or AST >400 iU/L) that was not due to viral hepatitis A-E, autoimmune hepatitis or poisoning. Control samples were obtained from children (aged <16 years) recruited to the Diagnosis and Management of Febrile Illness using RNA Personalised Molecular Signature Diagnosis (DIAMONDS), an ongoing multi-country study that aims to develop a molecular diagnostic test for the rapid diagnosis of severe infection and inflammatory diseases using personalised gene signatures (ISRCTN12394803). Ethical approval was given by London-Dulwich Research Ethics Committee (20/HRA/1714). Controls included healthy controls (n=13), children with PCR-confirmed adenoviral infection with normal transaminases (n= 12), and children with raised transaminases without adenoviral infection (n=33), recruited between 19 May 2020 to 8 January 2022.

### Viral PCR

**RNA extraction** was carried out using the Biomerieux Easymag generic protocol. 300ul of plasma or sera was extracted and eluted into 80 ul of water.

**AAV2 quantitative PCR** was performed to detect a 62bp amplicon of the AAV2 inverted terminal repeat region (ITR) as previously described^27^ using the forward ITR primer 5’-GGAACCCCTAGTGATGGAGTT-3’) and the reverse ITR primer 5’- CGGCCTCAGTGAGCGA-3’). The AAV2 ITR hydrolysis probe was labelled with fluorescein (6FAM) and quenched with Black Hole quencher (BHQ) 5’-[6FAM]- CACTCCCTCTCTGCGCGCTCG-[BHQ1]3’). AAV2 primers and probe were synthesised by Merck Life Sciences UK Limited, United Kingdom. Quantitative PCR analysis was performed using the ABI7500 Fast Real-Time PCR system (Applied Biosystems). LUNA Universal One-Step RT PCR kit (New England Biolabs) was used for the amplification and detection of the AAV2 ITR target. qPCR reactions were performed in a 20µl volume reaction (Luna Universal One-Step reaction mix, Luna WarmStart RT enzyme mix, 400nM forward and reverse primers, 200nM AAV2 ITR probe and 1-2.5µl of template DNA) as per manufacturer’s instructions. To quantify the number of copies, serial dilutions of plasmid containing the 62bp ITR product were used to generate a standard curve which was then used to interpolate the copy number of AAV2 copies in the samples. Wells with no template were used as negative controls. qPCR reactions were performed in triplicate. The qPCR program consisted of an initial reverse transcription step at 55 C for 10 minutes, an initial denaturation at 95 C for 1 minute followed by 45 cycles 95 C denaturation for 10 seconds and extension at 58 C for 1 minute. A qPCR detection limit of 31.9 was calculated as the threshold Ct value at the last dilution of DNA standards that were within the linear range.

The West of Scotland Specialist Virology Centre, NHS Greater Glasgow and Clyde conducted diagnostic real-time PCR with reverse transcription to detect HAdV, SARS-CoV-2-positive samples and other viral pathogens associated with hepatitis (e.g. Hepatitis A-E), following nucleic acid extraction utilizing the NucliSENS easyMAG and Roche MG96 platforms. HHV6^28^ and HAdV41^29^ were tested by qPCR as previously described using Invitrogen platinum qPCR mix (Cat no 11730-025) and Quanta Biosciences qPCR mix Mastermix (Cat.No. 733-1273) respectively on an ABI7500 and amplified for 40 cycles. A 6ul extract was amplified in a total reaction volume of 15ul.

### Measurement of antibody response to coronaviruses by electrochemiluminescence

IgG antibody titres were measured quantitatively against SARS-CoV-2 trimeric spike (S) protein, N-terminal domain (NTD), receptor binding domain (RBD) or nucleocapsid (N), and the human seasonal coronaviruses (HCoVs) 229E, OC43, NL63 and HKU1 using MSD V-PLEX COVID-19 Coronavirus Panel 2 (K15369) and Respiratory Panel 1 (K15365) kits. Multiplex Meso Scale Discovery electrochemiluminescence (MSD-ECL) assays were performed according to manufacturer instructions. Samples were diluted 1:5000 in diluent and added to the plates along with serially diluted reference standard (calibrator) and serology controls 1.1, 1.2 and 1.3. Plates were read using a MESO Sector S 600 plate reader. Data were generated by Methodological Mind software and analysed using MSD Discovery Workbench (v4.0). Results are expressed as MSD arbitrary units per ml (AU/ml).

### Metagenomic sequencing

Residual nucleic acid from 27 samples (9 patients with a combination of plasma, liver, faeces, rectal and throat/nose samples), 12 HAdV-positive and 13 healthy controls (control samples were either plasma or sera) underwent metagenomic next-generation sequencing at the CVR. Briefly, each nucleic acid sample was split in two library preparations, to improve the chances of detecting RNA and DNA viruses. The protocol applied for improved detection of RNA viruses included treatment with DNaseI (Ambion DNase I, ThermoFisher), ribosomal depletion (Ribo-Zero Plus rRNA Depletion Kit, Illumina), except for plasma samples, reverse transcription (SuperScript III, Invitrogen) and double-strand DNA synthesis (NEBNext® Ultra™ II Non-Directional RNA Second Strand Synthesis Module, NEB). The protocol applied to detect DNA viruses included partial removal of host DNA (NEBNext® Microbiome DNA Enrichment Kit, NEB). Following this, both sets of samples were used to prepare libraries using the KAPA LTP kit (Roche) with unique dual indices (NEBNext® Multiplex oligos for Illumina, NEB). The resulting libraries were pooled in equimolar amounts and sequenced using a NextSeq500 (Illumina) to obtain paired end reads using 150X150 cycles.

### Target enrichment sequencing

Following on the library preparation step described above, DNA and RNA derived libraries were pooled separately and were incubated with the VirCapSeq-VERT Capture Panel probes (Roche) following the manufacturer’s guidelines. The Roche VirCapSeq-VERT Capture Panel covers the genomes of 207 viral taxa known to infect vertebrates (including humans). Enriched DNA and RNA- derived libraries were further amplified using 14 PCR cycles, pooled and sequenced using a NextSeq500 (Illumina) to obtain paired end reads using 150X150 cycles.

### Bioinformatics analysis

Reads for each sample were first quality checked, Illumina adapters were trimmed using Trim Galore (https://github.com/FelixKrueger/TrimGalore), and reads were then mapped to the human genome using BWA-MEM (https://github.com/lh3/bwa). Only reads that did not map to the human genome were used for metagenomic analyses. Non-human reads were then *de novo* assembled using MetaSPAdes (https://github.com/ablab/spades) to generate contigs for each sample. Contigs were then compared against a protein database of all NCBI RefSeq organisms (including virus, bacteria, eukaryotes) with BLASTX using DIAMOND (https://github.com/bbuchfink/diamond). In addition, non-human reads for each sample were aligned to a small panel of HAdV NCBI RefSeq genomes (HAdV-A, B1, B2, C, D, E, F, 1, 2, 5, 7, 35, 54 as well as HAdV-F41).

The nine AAV2 near-complete genome contigs from the plasma samples were assembled and were compared with sequences in GenBank using BLASTN (nt database); each AAV2 had numerous close hits (exhibiting >95% similarity across 95% of the genome) with various existing AAV2 sequences, those most closely related were reported in a recent publication^25^ A representative range of AAV2 sequences were selected and combined with the AAV sequences *de novo* assembled here, aligned with MAFFT, the terminal ends of the alignment were trimmed off, and IQ-TREE 2 was used (TIM+F+R3 model) to infer a phylogenetic tree. For the single HAdV41 genome *de novo* assembled, all available HAdV41 complete genomes were downloaded from GenBank aligned with MAFFT, and IQ-TREE2 was used (K2P+R2 model) to infer a phylogenetic tree.

### HLA typing

High resolution typing for all HLA loci (HLA-A, B, C, DRB1, DRB3/4/5, DQA1, DQB1, DPA1 and DPB1) was performed using AllType™ FASTplex™ NGS Assay (One Lambda) run on an Illumina Mi-Seq platform. HLA typing was undertaken on 20 ISARIC consented patients. One patient was omitted from analysis as they were a sibling of another case. HLA types from 64 Scottish National Blood Transfusion Service apheresis platelet donors, self-identified as White British (n=15) or White Scottish (n=49) were used as control samples for comparison with patient HLA phenotype frequencies.

### Statistics

Differences between cases and control groups were calculated using Fisher’s Exact Test for categorical variables and Mann-Whitney test for continuous variables.

HLA analysis used the Bridging ImmunoGenomic Data-Analysis Workflow Gaps (BIGDAWG) R package to derive OR and corrected p values for individual HLA alleles.^30^ Bonferroni corrected p value significance threshold, adjusted for multiple comparisons (168 HLA alleles), was p < 3.0×10^−4^. Significance differences between groups for other comparisons were calculated using Mann-Whitney test or t-test as appropriate.

**Data and code availability:** available on request

### Funding

The work was funded by Public Health Scotland, the National Institute for Health Research (NIHR; award CO-CIN-01) and the Medical Research Council (MRC; grants MC UU 1201412, MC_PC_19059 & MC_PC_22004). DIAMONDS is funded by the European Union Horizon 2020 programme; grant 848196).

We wish to acknowledge the contribution of the participating children and their parents who agreed to participate in the ISARIC CCP-UK and DIAMONDS studies, and the research teams who recruited the patients. We also acknowledge the invaluable input of Eric J. Kremer from the Institut de Génétique Moléculaire de Montpellier, Université de Montpellier.

### Competing interest declaration

The authors have no competing interests

Supplementary Information is available for this paper.

Correspondence and requests for materials should be addressed to Professor Emma C Thomson.

Reprints and permissions information is available at www.nature.com/reprints.

## Data Availability

All data produced in the present study are available upon reasonable request to the authors, other than patient-specific data that cannot be released for reasons of confidentiality under the relevant ethical approvals.

## CONSORTIA

**ISARIC Comprehensive Clinical Characterisation Collaboration (ISARIC4C)**

J Kenneth Baillie, Malcolm G Semple, Gail Carson, Peter JM Openshaw, Jake Dunning, Laura Merson, Clark D Russell, Maria Zambon, Meera Chand, Richard S Tedder, Saye Khoo, Lance CW Turtle, Tom Solomon, Samreen Ijaz, Tom Fletcher, Massimo Palmarini, Antonia Ho, Emma C Thomson, Nicholas Price, Thushan de Silva, Chloe Donohue, Hayley Hardwick, Wilna Oosthuyzen, Miranda Odam, Primrose Chikowore, Sara Clohisey, Andrew Law, Lucy Norris, Sarah Tait, Murray Wham, Richard Clark, Audrey Coutts, Lorna Donnelly, Angie Fawkes, Tammy Gilchrist, Katarzyna Hafezi, Louise MacGillivray, Alan Maclean, Sarah McCafferty, Kirstie Morrice, Lee Murphy, Nicola Wrobel, Sarah E McDonald, Victoria Shaw, Katie A. Ahmed, Jane A Armstrong, Lauren Lett, Paul Henderson, Louisa Pollock, Shyla Kishore, Helen Brotherton, Lawrence Armstrong, Andrew Mitra, Anna Dall, Kristyna Bohmova, Sheena Logan, Louise Gannon, Ken Agwuh, Srikanth Chukkambotla, Ingrid DuRand, Duncan Fullerton, Sanjeev Garg, Clive Graham, Tassos Grammatikopoulos, Stuart Hartshorn, Luke Hodgson, Paul Jennings, George Koshy, Tamas Leiner, James Limb, Jeff Little, Sheena Logan, Elijah Matovu, Fiona McGill, Craig Morris, John Morrice, David Price, Henrik Reschreiter, Tim Reynolds, Paul Whittaker, Thomas Jordan, Rachel Tayler, Clare Irving, Katherine Jack, Maxine Ramsay, Margaret Millar, Barry Milligan, Naomi Hickey, Maggie Connon, Catriona Ward, Laura Beveridge, Susan MacFarlane, Karen Leitch, Claire Bell, Lauren Finlayson, Joy Dawson, Janie Candlish, Laura McGenily, Tara Roome, Cynthia Diaba, Jasmine Player, Natassia Powell, Ruth Howman, Sara Burling, Sharon Floyd, Sarah Farmer, Susie Ferguson, Susan Hope, Lucy Rubick, Rachel Swingler, Emma Collins, Collette Spencer, Amaryl Jones, Barbara Wilson, Diane Armstrong, Mark Birt, Holly Dickinson, Rosemary Harper, Darran Martin, Amy Roff, Sarah Mills.

**DIAMONDS Consortium**

PARTNER: Imperial College (Coordinating Centre) (UK) Chief investigator/DIAMONDS coordinator:

Michael Levin1

Principal and co-investigators (alphabetical order)1

Aubrey Cunnington; Jethro Herberg; Myrsini Kaforou; Victoria Wright

Section of Paediatric Infectious Diseases Research Group (alphabetical order)1

Evangelos Bellos; Claire Broderick; Samuel Channon-Wells; Samantha Cooray; Tisham De (database work package lead); Giselle D’Souza; Leire Estramiana Elorrieta; Diego Estrada-Rivadeneyra; Rachel Galassini (Clinical Trial Manager); Dominic Habgood-Coote; Shea Hamilton (Proteomics); Heather Jackson; James Kavanagh; Mahdi Moradi Marjaneh; Stephanie Menikou; Samuel Nichols; Ruud Nijman; Harsita Patel; Ivana Pennisi; Oliver Powell; Ruth Reid; Priyen Shah; Ortensia Vito; Elizabeth Whittaker; Clare Wilson; Rebecca Womersley

Recruitment team at Imperial College Healthcare NHS Trust, London (alphabetical order)2 Amina Abdulla; Sarah Darnell; Sobia Mustafa

Engineering Team

Pantelis Georgiou3 (engineering lead); Jesus-Rodriguez Manzano4; Nicolas Moser3; Ivana Pennisi1

1 Section of Paediatric Infectious Disease, Imperial College London, Norfolk Place, London W2 1PG, UK

2 Children’s Clinical Research Unit, St Mary’s Hospital, Praed Street, London W2 1NY, UK

3 Imperial College London, Department of Electrical and Electronic Engineering, South Kensington Campus, London, SW7 2AZ, UK

4 Imperial College London, Department of Infectious Disease, Section of Adult Infectious Disease, Hammersmith Campus, London, W12 0NN, UK

UK Non-Consortium Clinical Recruiting Sites

Evelina London Children’s Hospital, Guy’s and St Thomas’ NHS Foundation Trust; King’s College London [combined]

Michael Carter1,2 (principal investigator); Shane Tibby1,2 (co-investigator)

Recruitment team (alphabetical order): Jonathan Cohen1; Francesca Davis1; Julia Kenny1; Paul Wellman1; Marie White1

Laboratory team (alphabetical order): Matthew Fish3; Aislinn Jennings4; Manu Shankar-Hari3,4

1 Evelina London Children’s Hospital, Guy’s and St Thomas’ NHS Foundation Trust, London, UK

2 Department of Women and Children’s Health, School of Life Course Sciences, King’s College London, UK

3 Department of Infectious Diseases, School of Immunology and Microbial Sciences, King’s College London, London, UK

4 Department of Intensive Care Medicine, Guy’s and St Thomas’ NHS Foundation Trust, London, UK

University Hospitals Sussex

Katy Fidler1 (principal investigator); Dan Agranoff2 (co-investigator) Recruitment team; Vivien Richmond1,3, Mathhew Seal2

1 Royal Alexandra Children’s Hospital, University Hospitals Sussex, Brighton, UK

2 Dept of Infectious Diseases, University Hospitals Sussex, Brighton, UK

3 Research Nurse team, University Hospitals Sussex, Brighton, UK

University Hospital Southampton NHS Foundation Trust

Saul Faust1 (principal investigator); Dan Owen1 (co-investigator);

Recruitment team; Ruth Ensom2; Sarah McKay2; Diana Mondo3, Mariya Shaji3; Rachel Schranz3 (alphabetical order)

1 NIHR Southampton Clinical Research Facility, University Hospital Southampton NHS Foundation Trust and University of Southampton, UK

2 NIHR Southampton Clinical Research Facility, University Hospital Southampton NHS Foundation Trust, UK

3 Department of R&D, University Hospital Southampton NHS Foundation Trust, UK

Barts Health NHS Trust

Prita Rughnani1, 2, 3 (principal investigator 2020-2021); Amutha Anpananthar1, 2, 3 (principal investigator 2021-to date); Susan Liebeschuetz2 (co-investigator), Anna Riddell1 (co-investigator)

Recruitment team; Nosheen Khalid1, 3,Ivone Lancoma Malcolm, Teresa Simagan3 (alphabetical order)

1 Royal London Hospital, Whitechapel Rd, London E1 1FR, UK

2 Newham University Hospital, Glen Rd, London E13 8SL, UK

3 Whipps Cross University Hospital, Whipps Cross Road, London, E11 1NR, UK

Great Ormond Street Hospital for Children NHS Foundation Trust

Mark Peters1,2 (principal investigator); Alasdair Bamford1,2 (co-investigator) Recruitment team; Lauran O’Neill1

1 Great Ormond Street Hospital, London, WC1N 3JH, UK

2 UCL Great Ormond St Institute of Child Health, WC1N 1EH, UK

Cambridge University Hospitals NHS Foundation Trust Nazima Pathan1,2 (principal investigator)

Recruitment team; Esther Daubney1, Deborah White1 (alphabetical order)

1 Addenbrooke’s Hospital, Hills Road, Cambridge CB2 0QQ, UK

2 Department of Paediatrics, University of Cambridge, Cambridge CB2 0QQ, UK

University College London Hospitals NHS Foundation Trust

Melissa Heightman1 (principal investigator); Sarah Eisen1 (co-investigator)

Recruitment team; Terry Segal1, Lucy Wellings1 (alphabetical order)

1 University College London Hospital, Euston Road, London NW1 2BU, UK

St George’s University Hospitals NHS Foundation Trust Simon B Drysdale1 (principal investigator)

Recruitment team; Nicole Branch1, Lisa Hamzah1, Heather Jarman1 (alphabetical order) 1 St George’s Hospital, Blackshaw Road, London SW17 0QT, UK

Lewisham and Greenwich NHS Trust Maggie Nyirenda1, 2,(principal investigator)

Recruitment team Lisa Capozzi1, Emma Gardiner1 (alphabetical order)

1 University Hospital Lewisham, London SE13 6LH, UK

2 Queen Elizabeth Hospital Greenwich, London SE18 4QH, UK

Liverpool University Hospitals NHS Foundation Trust

Robert Moots1 (principal investigator); Magda Nasher2 (principal investigator) Recruitment team; Anita Hanson2; Michelle Linforth1

1 Aintree University Hospital, Lower Lane, Liverpool L9 7AL, UK

2 Royal Liverpool Hospital, Prescot St, Liverpool L7 8XP, UK

Leeds Teaching Hospitals NHS Trust Sean O’Riordan1 (principal investigator) Recruitment team; Donna Ellis1

1Leeds Children’s Hospital, Leeds LS1 3EX, UK

King’s College Hospital NHS Foundation Trust Akash Deep1 (principal investigator) Recruitment team; Ivan Caro1

1 Kings College Hospital, Denmark Hill, London SE5 9RS, UK

Sheffield Children’s NHS Foundation Trust Fiona Shackley 1 (principal investigator);

Recruitment team; Arianna Bellini,1 Stuart Gormley1 (alphabetical order) 1Sheffield Children’s Hospital, Broomhall, Sheffield S10 2TH, UK

University Hospitals of Leicester NHS Foundation Trust Samira Neshat1 (principal investigator)

1Leicester General Hospital, Leicester LE1 5WW, UK

Birmingham Women’s and Children’s Hospital NHS Foundation Trust Barnaby J Scholefield1 (principal investigator)

Recruitment team; Ceri Robbins1, Helen Winmill1 (alphabetical order)

1 Birmingham Children’s Hospital, Steelhouse Lane, Birmingham B4 6NH, UK PARTNER: University of Oxford (UK)

Children’s Hospital, John Radcliffe Hospital, Oxford

Principal Investigator Stéphane C. Paulus1,2,3 Co-Principal Investigator

Andrew J. Pollard1,2,3,4

Co-investigators

Mark Anthony1 (neonates)

Recruitment team

Sarah Hopton1, Danielle Miller1, Zoe Oliver1, Sally Beer1, Bryony Ward1

1 John Radcliffe Hospital, Oxford University Hospitals NHS Foundation Trust, Oxford, UK

2 Department of Paediatrics, University of Oxford, UK

3 Oxford Vaccine Group, University of Oxford, UK 4NIHR Oxford Biomedical Research Centre, Oxford, UK

University of Oxford, Nepal Site Principal Investigator

Shrijana Shrestha1

Co-Principal Investigator Andrew J Pollard2,3

Nepal Research Team Meeru Gurung1

Puja Amatya1 Bhishma Pokhrel1

Sanjeev Man Bijukchhe1 Oxford Research Team Tim Lubinda2

Sarah Kelly2 Peter O’Reilly2

1 Paediatric Research Unit, Patan Academy of Health Sciences, Kathmandu, Nepal.

2 Oxford Vaccine Group, Department of Paediatrics, University of Oxford, Oxford, United Kingdom.

3 NIHR Oxford Biomedical Research Centre, Oxford, United Kingdom.

PARTNER: SERGAS (Spain)

Principal Investigators Federico Martinón-Torres1 Antonio Salas1,2

GENVIP RESEARCH GROUP (in alphabetical order):

Fernando Álvez González1, Xabier Bello1,2, Mirian Ben García1, Sandra Carnota1, Miriam Cebey- López1, María José Curras-Tuala1,2, Carlos Durán Suárez1, Luisa García Vicente1, Alberto Gómez- Carballa1,2, Jose Gómez Rial1, Pilar Leboráns Iglesias1, Federico Martinón-Torres1, Nazareth Martinón-Torres1, José María Martinón Sánchez1, Belén Mosquera Pérez1, Jacobo Pardo-Seco1,2, Lidia Piñeiro Rodríguez1, Sara Pischedda1,2, Sara Rey Vázquez1, Irene Rivero Calle1, Carmen Rodríguez-Tenreiro1, Lorenzo Redondo-Collazo1, Miguel Sadiki Ora1, Antonio Salas1,2, Sonia Serén Fernández1, Cristina Serén Trasorras1, Marisol Vilas Iglesias1.

1 Translational Pediatrics and Infectious Diseases, Pediatrics Department, Hospital Clínico Universitario de Santiago, Santiago de Compostela, Spain, and GENVIP Research Group (www.genvip.org), Instituto de Investigación Sanitaria de Santiago, Universidad de Santiago de Compostela, Galicia, Spain.

2 Unidade de Xenética, Departamento de Anatomía Patolóxica e Ciencias Forenses, Instituto de Ciencias Forenses, Facultade de Medicina, Universidade de Santiago de Compostela, and GenPop Research Group, Instituto de Investigaciones Sanitarias (IDIS), Hospital Clínico Universitario de Santiago, Galicia, Spain

3 Fundación Pública Galega de Medicina Xenómica, Servizo Galego de Saúde (SERGAS), Instituto de Investigaciones Sanitarias (IDIS), and Grupo de Medicina Xenómica, Centro de Investigación Biomédica en Red de Enfermedades Raras (CIBERER), Universidade de Santiago de Compostela (USC), Santiago de Compostela, Spain

PARTNER: Liverpool (UK) Principal Investigators Enitan D Carrol1,2,

Research Group (in alphabetical order):

Elizabeth Cocklin1, Aakash Khanijau1, Rebecca Lenihan1, Nadia Lewis-Burke1

Karen Newall4, Sam Romaine1,

1 Department of Clinical Infection, Microbiology and Immunology, University of Liverpool Institute of Infection, Veterinary and Ecological Sciences, Liverpool, England

2 Alder Hey Children’s Hospital, Department of Infectious Diseases, Eaton Road, Liverpool, L12 2AP

3 4Alder Hey Children’s Hospital, Clinical Research Business Unit, Eaton Road, Liverpool, L12 2AP

PARTNER: NATIONAL AND KAPODISTRIAN UNIVERSITY OF ATHENS (Greece)

Principal Investigator: Maria Tsolia1 Co-Investigator: Irini Eleftheriou1

PID Unit: Nikos Spyridis1, Maria Tambouratzi1 Pediatric Rheumatology Unit: Despoina Maritsi1 Lab: Antonios Marmarinos1, Marietta Xagorari1

Recruitment teams:

Adult COVID19- Infectious Diseases: Lourida Panagiota, Pefanis Aggelos2 Adult COVID19: Akinosoglou Karolina, Gogos Charalambos, Maragos Markos3

Adult Inflammatory Diseases-Oncology: Voulgarelis Michalis, Stergiou Ioanna4

1 2nd Department of Pediatrics, National and Kapodistrian University of Athens (NKUA), Children’s Hospital “P, and A. Kyriakou”, Athens, Greece

2 1st Department of Infectious Diseases, General Hospital “Sotiria”

3 Pathology Department, University of Patras, General Hospital “Panagia i Voithia”

4 Pathophysiology Department, Medical Faculty, National and Kapodistrian University of Athens (NKUA), General Hospital “Laiko”

Newcastle upon Tyne Hospitals NHS Foundation Trust and Newcastle University (UK) combined

Principal Investigator:

Marieke Emonts 1,2,3 (all activities)

Co-investigators

Emma Lim2,3,6 (all activities) John Isaacs1 (adult inflammatory)

Recruitment team (alphabetical), datamanagers, and GNCH Research unit:

Kathryn Bell4, Stephen Crulley4, Daniel Fabian4, Evelyn Thomson4, Diane Wallia4, Caroline Miller4, Ashley Bell4

PhD Students/medical staff DIAMONDS

Fabian J.S. van der Velden1,2 (all activities), Geoff Shenton7 (oncology), Ashley Price8,9 (Adult COVID)

Students

Owen Treloar 1,2 (quality control, data management and analysis) Daisy Thomas1,2 (recruitment)

Author Affiliations:

1 Translational and Clinical Research Institute, Newcastle University, Newcastle upon Tyne UK

2 Great North Children’s Hospital, Paediatric Immunology, Infectious Diseases & Allergy, Newcastle upon Tyne Hospitals NHS Foundation Trust, Newcastle upon Tyne, United Kingdom.

3 NIHR Newcastle Biomedical Research Centre based at Newcastle upon Tyne Hospitals NHS Trust and Newcastle University, Westgate Rd, Newcastle upon Tyne NE4 5PL, United Kingdom

4 Great North Children’s Hospital, Research Unit, Newcastle upon Tyne Hospitals NHS Foundation Trust, Newcastle upon Tyne, United Kingdom.

6 Population Health Sciences Institute, Newcastle University, Newcastle upon Tyne, UK

7 Great North Children’s Hospital, Paediatric Oncology, Newcastle upon Tyne Hospitals NHS Foundation Trust, Newcastle upon Tyne, United Kingdom.

8 Department of Infection & Tropical Medicine, Newcastle upon Tyne Hospitals NHS Foundation Trust, Newcastle upon Tyne, United Kingdom

9 NIHR Newcastle In Vitro Diagnostics Co-operative (Newcastle MIC), Newcastle upon Tyne, United Kingdom.

Servicio Madrileño de Salud (SERMAS) - Fundación Biomédica del Hospital Universitario 12 de Octubre (FIB-H12O) (Spain)

Principal Investigators

Pablo Rojo1 3

Cristina Epalza 1,2 SERMAS/FIB-H120 team:

Serena Villaverde 1,, Sonia Márquez2, Manuel Gijón 2, Fátima Machín2, Laura Cabello2, Irene Hernández2, Lourdes Gutiérrez2, Ángela Manzanares 1

Author Affiliations:

1 Servicio Madrileño de Salud (SERMAS),Pediatric Infectious Diseases Unit, Department of Pediatrics, Hospital Universitario 12 de Octubre, Madrid, Spain

2 Fundación Biomédica del Hospital Universitario 12 de Octubre (FIB-H12O), Unidad Pediátrica de Investigación y Ensayos Clínicos (UPIC), Hospital Universitario 12 de Octubre, Instituto de Investigación Sanitaria Hospital 12 de Octubre (i+12), Madrid, Spain.

3 Universidad Complutense de Madrid, Faculty of Medicine, Department of Pediatrics, Madrid, Spain.

Amsterdam University Medical Center (Amsterdam UMC), University of Amsterdam

Principal Investigator:

T.W. (Taco) Kuijpers MD PhD 1,2 (all activities)

Co-investigators

1. M. (Martijn) van de Kuip MD PhD 1 (infectious disease)

A.M. (Marceline) van Furth MD PhD 1 (infectious disease)

J.M. (Merlijn) van den Berg MD PhD 1 (inflammatory disease)

Hospital Team (all activities):

Giske Biesbroek MD PhD 1, Floris Verkuil MD (PhD student) 1, Carlijn (C.W.) van der Zee MD (start 1/8/2022, PhD student) 1

Recruitment:

Dasja Pajkrt MD PhD 1, Michael Boele van Hensbroek MD PhD 1, Dieneke Schonenberg MD 1, Mariken Gruppen MD 1, Sietse Nagelkerke MD PhD 1,2, medical students

Laboratory Team:

Machiel H Jansen 1, Ines Goetschalckx (PhD student) 2

Author Affiliations:

1 Amsterdam UMC, Emma Children’s Hospital, Dept of Pediatric Immunology, Rheumatology and Infectious Disease, University of Amsterdam, The Netherlands

2 Sanquin, Dept of Molecular Hematology, University Medical Center, Amsterdam, The Netherlands

Bambino Gesù Children’s Hospital (Rome-Italy) Principal Investigator

Lorenza Romani 1 Maia De Luca 1

Recruitment Team Sara Chiurchiù 1 Martina Di Giuseppe 1

Affiliation

1 Infectious Disease Unit, Academic Department of Pediatrics, Bambino Gesù Children’s Hospital, IRCCS, Rome 00165, Italy

ERASMUS MC-Sophia Children’s Hospital Principal Investigator

Clementien L. Vermont²

Research group

Henriëtte A. Moll¹, Dorine M. Borensztajn¹, Nienke N. Hagedoorn, Chantal Tan ¹, Joany Zachariasse ¹, Medical students ¹

Additional investigator W Dik 3

1 Erasmus MC-Sophia Children’s Hospital, Department of General Paediatrics, Rotterdam, the Netherlands

2 Erasmus MC-Sophia Children’s Hospital, Department of Paediatric Infectious Diseases & Immunology, Rotterdam, the Netherlands

3 Erasmus MC, Department of immunology, Rotterdam, the Netherlands

TAIWAN

Ching-Fen (Kitty), Shen

Division of Infectious Disease, Department of Pediatrics, National Cheng Kung University Tainan, Taiwan

Riga Stradins University (Riga, Latvia)

Principal Investigator:

Dace Zavadska 1,2 (all activities)

Co-investigators

Sniedze Laivacuma 1,3 (adult cohorts)

Recruitment team:

Aleksandra Rudzate 1,2, Diana Stoldere 1,2, Arta Barzdina 1,2, Elza Barzdina 1,2, Sniedze Laivacuma1,3, Monta Madelane 1,3

Laboratory

Dagne Gravele 2, Dace Svile2

Author Affiliations:

1 Riga Stradins University, Riga, Latvia

2 Children clinical university hospital, Riga, Latvia

3 Riga East clinical university hospital, Riga, Latvia

Assistance Publique - Hôpitaux de Paris Principal Investigator:

Romain Basmaci 1,2

Co-investigator: Noémie Lachaume 1

Recruitment team:

Pauline Bories 1, Raja Ben Tkhayat 1, Laura Chériaux 1, Juraté Davoust 1, Kim-Thanh Ong 1, Marie Cotillon 1, Thibault de Groc 1, Sébastien Le 1, Nathalie Vergnault 1, Hélène Sée 1, Laure Cohen 1, Alice de Tugny 1, Nevena Danekova 1

Author Affiliations:

1 Service de Pédiatrie-Urgences, AP-HP, Hôpital Louis-Mourier, F-92700 Colombes, France ² Université Paris Cité, Inserm, IAME, F-75018 Paris, France

BioMérieux

Principal Investigator:

Marine Mommert-Tripon Co-investigator:

Karen Brengel-Pesce

Author Affiliations:

bioMérieux - Open Innovation & Partnerships Department, Lyon, France

University Medical Centre Ljubljana, Slovenia

Principal Investigator: Marko Pokorn 1,2,3 Co-Investigator: Mojca Kolnik2

Research Group (in alphabetical order):

Tadej Avčin2,3, Tanja Avramoska2, Natalija Bahovec1, Petra Bogovič1, Lidija Kitanovski2,3, Mirijam Nahtigal1, Lea Papst1, Tina Plankar Srovin1, Franc Strle1,2, Anja Srpčič2, Katarina Vincek1.

Affiliations:

1. Department of Infectious diseases, University Medical Centre Ljubljana, Slovenia

2. University Children’s Hospital, University Medical Centre Ljubljana, Slovenia

3. Faculty of Medicine, University of Ljubljana, Slovenia

4. Centre for Clinical research, University Medical Centre Ljubljana

University Medical Center Utrecht, Utrecht, The Netherlands Principal Investigator

Michiel van der Flier1,5 (Pediatric Infectious Diseases and Immunology)

Co-investigators

Wim J.E. Tissing5 (Pediatric Oncology)

Roelie M. Wösten-van Asperen2 (Pediatric Intensive Care Unit) Sebastiaan J Vastert3 (Pediatric Rheumatology)

Daniel C Vijlbrief4 (Pediatric Neonatal Intensive Care)

Louis J. Bont1,5 (Pediatric Infectious Diseases and Immunology) Tom F.W. Wolfs 1,5 (Pediatric Infectious Diseases and Immunology)

PhD student

Coco R. Beudeker1,5 (Pediatric Infectious Diseases and Immunology)

Affiliations:

1.Pediatric Infectious Diseases and Immunology, 2. Pediatric Intensive Care Unit 3. Pediatric Rheumatology 4. Pediatric Neonatal Intensive Care, Wilhelmina Children’s Hospital, University Medical Center Utrecht, Utrecht, The Netherlands

1 5. Princess Maxima Center for Pediatric Oncology, Utrecht, The Netherlands

PARTNER: University of Bern, Inselspital, Bern University Hospital, University of Bern (Switzerland)

Principal Investigators (alphabetical) Philipp Agyeman1

Luregn Schlapbach2,3

Co-Investigator Christoph Aebi1

Recruitment team

Mariama Usman1, Stefanie Schlüchter1, Verena Wyss1, Nina Schöbi1, Elisa Zimmermann2 PhD, Marion Meier2, Kathrin Weber2

1 Department of Pediatrics, Inselspital, Bern University Hospital, University of Bern, Switzerland

2 Department of Intensive Care and Neonatology, and Children’s Research Center, University Children’s Hospital Zurich, Zurich, Switzerland

3 Child Health Research Centre, The University of Queensland, Brisbane, Australia

Swiss Pediatric Sepsis Study group

Philipp Agyeman, MD 1, Luregn J Schlapbach, MD, FCICM 2,3, Eric Giannoni, MD 4,5, Martin Stocker, MD 6, Klara M Posfay-Barbe, MD 7, Ulrich Heininger, MD 8, Sara Bernhard-Stirnemann, MD 9, Anita Niederer-Loher, MD 10, Christian Kahlert, MD 10, Giancarlo Natalucci, MD 11, Christa Relly, MD 12, Thomas Riedel, MD 13, Christoph Aebi, MD 1, Christoph Berger, MD 12

Affiliations:

1 Department of Pediatrics, Inselspital, Bern University Hospital, University of Bern, Switzerland

2 Department of Intensive Care and Neonatology, and Children’s Research Center, University Children’s Hospital Zurich, Zurich, Switzerland

3 Child Health Research Centre, The University of Queensland, Brisbane, Australia

4 Clinic of Neonatology, Department Mother-Woman-Child, Lausanne University Hospital and University of Lausanne, Switzerland

5 Infectious Diseases Service, Department of Medicine, Lausanne University Hospital and University of Lausanne, Switzerland

6 Department of Pediatrics, Children’s Hospital Lucerne, Lucerne, Switzerland

7 Pediatric Infectious Diseases Unit, Children’s Hospital of Geneva, University Hospitals of Geneva, Geneva, Switzerland

8 Infectious Diseases and Vaccinology, University of Basel Children’s Hospital, Basel, Switzerland

9 Children’s Hospital Aarau, Aarau, Switzerland

10 Division of Infectious Diseases and Hospital Epidemiology, Children’s Hospital of Eastern Switzerland St. Gallen, St. Gallen, Switzerland

11 Department of Neonatology, University Hospital Zurich, Zurich, Switzerland

12 Division of Infectious Diseases and Hospital Epidemiology, and Children’s Research Center, University Children’s Hospital Zurich, Switzerland

13 Children’s Hospital Chur, Chur, Switzerland

Micropathology Ltd (UK)

Micropathology Ltd, The Venture Center, University of Warwick Science Park, Sir William Lyons Road, Coventry, CV4 7EZ

Principle Investigator; Prof Colin Fink

Co Investigators: Marie Voice, Leo Calvo-Bado, Michael Steele, Jennifer Holden Research group: Benjamin Evans, Jake Stevens, Peter Matthews, Kyle Billing

Medical University of Graz, Austria (MUG)

Principal Investigator:

Werner Zenz1 (all activities)

Co-investigators (in alphabetical order):

Alexander Binder1 (grant application)

Benno Kohlmaier1 (study design, recruitment) Daniela S. Kohlfürst1 (study design)

Nina A. Schweintzger1 (all activities) Christoph Zurl1 (study design, recruitment)

Recruitment team, data managers, laboratory work (in alphabetical order):

Susanne Hösele1, Manuel Leitner1, Lena Pölz1, Alexandra Rusu1, Glorija Rajic1, Bianca Stoiser1, Martina Strempfl1, Manfred G. Sagmeister1

Clinical recruitment partners (in alphabetical order):

Sebastian Bauchinger1, Martin Benesch3, Astrid Ceolotto1, Ernst Eber2, Siegfried Gallistl1, Harald Haidl1, Almuthe Hauer1, Christa Hude1, Andreas Kapper7, Markus Keldorfer5, Sabine Löffler5, Tobias Niedrist6, Heidemarie Pilch5, Andreas Pfleger2, Klaus Pfurtscheller4, Siegfried Rödl4, Andrea Skrabl-Baumgartner1, Volker Strenger3, Elmar Wallner7

Author Affiliations:

1 Department of Pediatrics and Adolescent Medicine, Division of General Pediatrics, Medical University of Graz, Graz, Austria

2 Department of Pediatric Pulmonology, Medical University of Graz, Graz, Austria

3 Department of Pediatric Hematooncology, Medical University of Graz, Graz, Austria

4 Paediatric Intensive Care Unit, Medical University of Graz, Graz, Austria

5 University Clinic of Pediatrics and Adolescent Medicine Graz, Medical University Graz, Graz, Austria

6 Clinical Institute of Medical and Chemical Laboratory Diagnostics, Medical University Graz, Graz, Austria

7 Department of Internal Medicine, State Hospital Graz II, Location West, Graz, Austria

SkylineDX

Principle investigator: Dennie Tempel 1

Co-investigators: Danielle van Keulen1, Annelieke M Strijbosch 1, Author affiliations:

1 SkylineDx, Rotterdam, The Netherlands

Project partner BBMRI-ERIC Maike K. Tauchert

Author affiliation:

Biobanking and BioMolecular Resources Research Infrastructure - European Research Infrastructure Consortium (BBMRI-ERIC), Neue Stiftingtalstrasse 2/B/6, 8010, Graz, Austria

LMU Munich Partner (Germany) Principal Investigator:

Ulrich von Both1,2 MD, FRCPCH (all activities)

Research group:

Laura Kolberg¹ MSc (all activities)

Patricia Schmied¹ (Study physician), Irene Alba-Alejandre3 MD (Study physician)

Clinical recruitment partners (in alphabetical order):

Katharina Danhauser, MD6, Nikolaus Haas, MD11, Florian Hoffmann, MD10, Matthias Griese, MD7, Tobias Feuchtinger, MD5, Sabrina Juranek, MD4, Matthias Kappler, MD7, Eberhard Lurz, MD8, Esther Maier, MD4, Karl Reiter, MD10, Carola Schoen, MD10, Sebastian Schroepf, MD9

Author Affiliations:

1 Division of Pediatric Infectious Diseases, Department of Pediatrics, Dr. von Hauner Children’s Hospital, University Hospital, LMU Munich, Munich, Germany

2 German Center for Infection Research (DZIF), Partner Site Munich, Munich, Germany

3 Department of Gynecology and Obstetrics, University Hospital, LMU Munich, Munich, Germany

4 Division of General Pediatrics, Department of Pediatrics, Dr. von Hauner Children’s Hospital, University Hospital, LMU Munich, Munich, Germany

5 Division of Pediatric Haematology & Oncology, Department of Pediatrics, Dr. von Hauner Children’s Hospital, University Hospital, LMU Munich, Munich, Germany

6 Division of Pediatric Rheumatology, Department of Pediatrics, Dr. von Hauner Children’s Hospital, University Hospital, LMU Munich, Munich, Germany

7 Division of Pediatric Pulmonology, Department of Pediatrics, Dr. von Hauner Children’s Hospital, University Hospital, LMU Munich, Munich, Germany

8 Division of Pediatric Gastroenterology, Department of Pediatrics, Dr. von Hauner Children’s Hospital, University Hospital, LMU Munich, Munich, Germany

9 Neonatal Intensive Care Unit, Department of Pediatrics, Dr. von Hauner Children’s Hospital, University Hospital, LMU Munich, Munich, Germany

10 Paediatric Intensive Care Unit, Department of Pediatrics, Dr. von Hauner Children’s Hospital, University Hospital, LMU Munich, Munich, Germany

11 Department of Pediatric Cardiology and Pediatric Intensive Care, University Hospital, LMU Munich, Germany

London School of Hygiene and Tropical Medicine (LSHTM) Principal Investigator: Shunmay Yeung 1,2,3

Research group:

Manuel Dewez1 David Bath3, Elizabeth Fitchett1, Fiona Cresswell1

1. Clinical Research Department, Faculty of Infectious and Tropical Disease, London School of Hygiene and Tropical Medicine, London

2. Department of Paediatrics, St. Mary’s Imperial College Hospital, London

3. Department of Global Health and Development, Faculty of Public Health and Policy, London School of Hygiene and Tropical Medicine, London

---

**Supplementary Table 1.** Characteristics of cases and controls.

|  | **Cases**<br>(n=9) | **Controls** |  |  |  |  |  |
| --- | --- | --- | --- | --- | --- | --- | --- |
|  |  | Healthy<br>(n=13) | P<br>value* | HAdV<br>infection with<br>normal<br>transaminases<br>(N=12) | P<br>value* | Elevated<br>transaminases<br>with no<br>HAdV<br>infection<br>(n=33) | P<br>value* |
| Sex - male | 4<br>(44.4) | 10 (76.9) | 0.187 | 7 (58.3) | 0.67 | 18 (52.9) | 0.71 |
| Age (years) | 3.9<br>(3.4-5.1) | 4.1 (3.6-4.9) | 0.89 | 1.6 (1.1-3.3) | <0.001 | 9.2 (6.7-13.6) | 0.001 |
| Recruitment<br>period | 14<br>March<br>2 - 4<br>April<br>2022 | 6 Nov 2020 -<br>6 Jul 2021 | - | 22 May 2020<br>- 22 Dec 2021 | - | 9 Sep 2020 - 8<br>Jan 2022 | - |

\*Fisher's exact or Mann-Whitney test

**Supplementary Table 2.** Clinical laboratory results of the nine non-A-E hepatitis cases in Scottish children.

| Patient |  | 1 | 2 | 3 | 4 | 5 | 6 | 7 | 8 | 9 |
| --- | --- | --- | --- | --- | --- | --- | --- | --- | --- | --- |
| **HAdV PCR** | **Blood** | Negative | Negative | Negative | Negative | Negative | Negative | Negative | Negative | Negative |
|  | **Throat/nose** | NT | Negative | Positive (Ct 38) | Negative | Negative | Negative | Negative | Positive (Ct 30) | NT |
|  | **Stool** | Negative | Negative | Negative | Negative | Positive (Ct 31) | Positive (Ct 34) | Negative | Positive (Ct 31) | NT |
| **HAdV 41 PCR** | **Blood** | Negative | Negative | Positive (Ct 36) | Negative | Negative | Negative | Negative | Negative | Positive (Ct 37) |
| **Hepatitis A** | **IgM** | Negative | Negative | Negative | Negative | Negative | Negative | Negative | Negative | Negative |
|  | **PCR** | Negative | Negative | Negative | Negative | Negative | NT | NT | NT | NT |
| **Hepatitis B** | **sAg** | Negative | Negative | Negative | Negative | Negative | Negative | Negative | Negative | Negative |
| **Hepatitis C** | **PCR** | Negative | NT (IgG negative) | Negative | Negative | Negative | Negative | NT | NT | Negative |
| **Hepatitis E** | **IgM** | Negative | Negative | Negative | Negative | Negative | Negative | Negative | Negative | Negative |
|  | **PCR** | Negative | Negative | Negative | Negative | Negative | NT | NT | NT | NT |
| **Parvovirus B19** | **IgM** | Negative | NT | Negative | Negative | Negative | Negative | Negative | Negative | NT |
|  | **PCR** | NT | Negative | NT | NT | NT | NT | NT | NT | NT |
| **CMV** | **IgM** | Negative | Negative | Negative | Negative | Negative | Negative | Negative | Negative | Negative |
| **EBV** | **IgM** | Negative | Negative | Negative | Negative | Negative | Negative | Negative | Negative | Negative |
| **Enterovirus PCR** | **Blood** | Negative | Negative | Negative | Negative | Negative | NT | NT | Negative | NT |
| **HSV PCR** | **Blood** | Negative | Negative | Negative | Negative | Negative | Negative | Negative | Negative | Negative |
| **HIV Ag/Ab** | **Blood** | Negative | Negative | NT | Negative | Negative | Negative | Negative | Negative | NT |
| **HHV6/7** | **Blood PCR** | Negative | Negative | Negative | Negative | Negative | Negative | Negative | Negative | Negative |
| **Respiratory viral PCR\*** | **Throat/ nose** | NT | Negative | hCoV-NL63 | Negative | Paraflu 3 | Paraflu 2 Rhino/EV | Negative | Rhino/EV (Ct 36) | NT |
| **SARS-CoV-2 PCR** | **Throat/ nose** | Negative | Negative | Negative | Negative | Positive | Positive | Negative | Negative | Negative |
| **Liver** | **CMV, EBV, HSV, HHV6/7 PCR** | Positive HHV6 PCR+ (Ct 33) |  | Negative | Positive HHV6 PCR (Ct 36) |  |  |  |  | Negative |
| **Other sample** |  |  |  |  |  |  |  | Stool Norovirus PCR Positive |  |  |

Abbreviations – PCR, polymerase chain reaction; CMV, cytomegalovirus; EBV, Epstein-Barr Virus;
HSV, Herpes Simplex virus, HIV, Human Immunodeficiency Virus; HHV, Human herpesvirus; PCR,
polymerase chain reaction; NT, not tested

**Supplementary Table 3.**
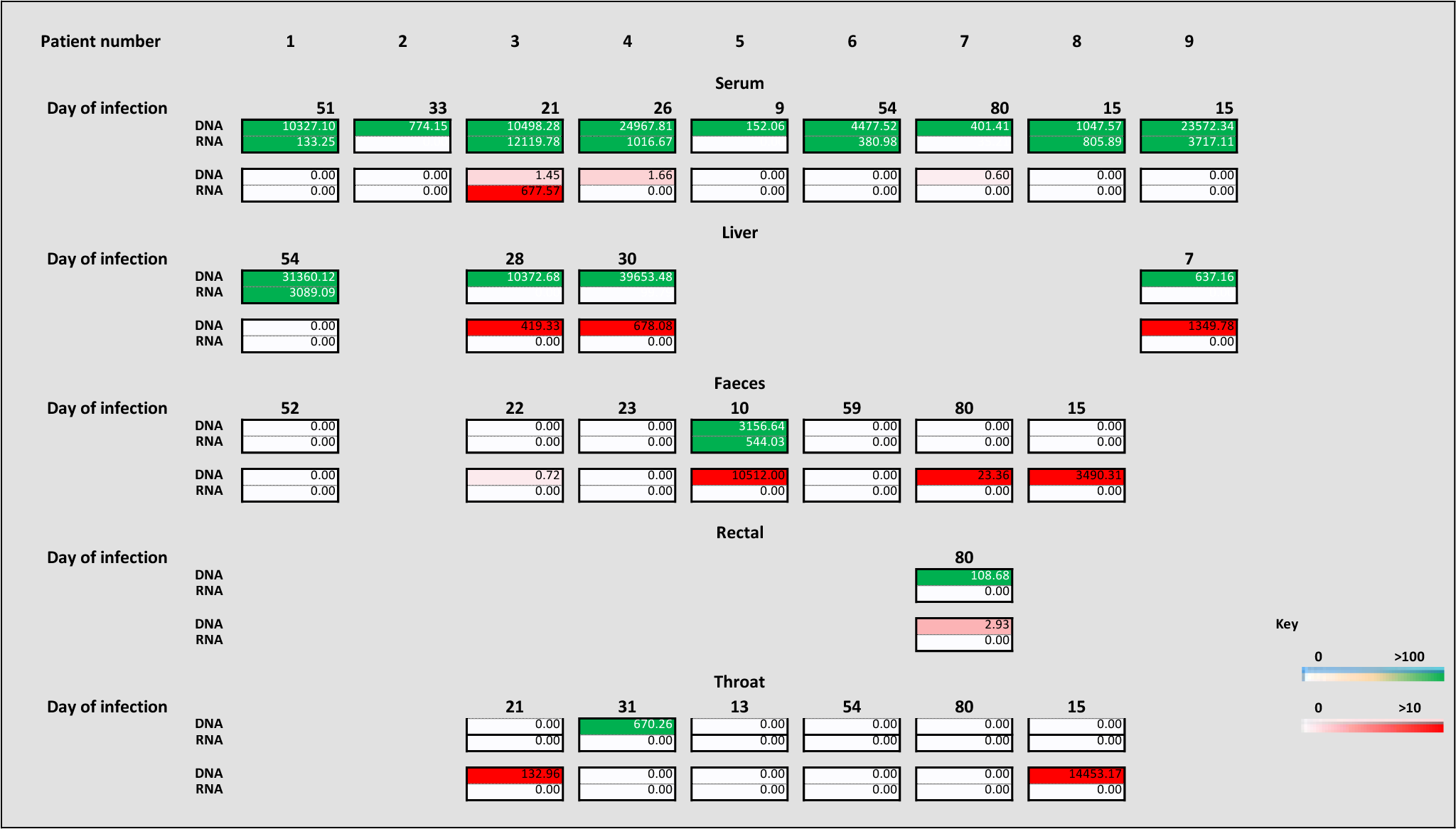
Read counts per million of AAV2 and HAdV in cases following target enrichment sequencing.

**Supplementary Table 4.**
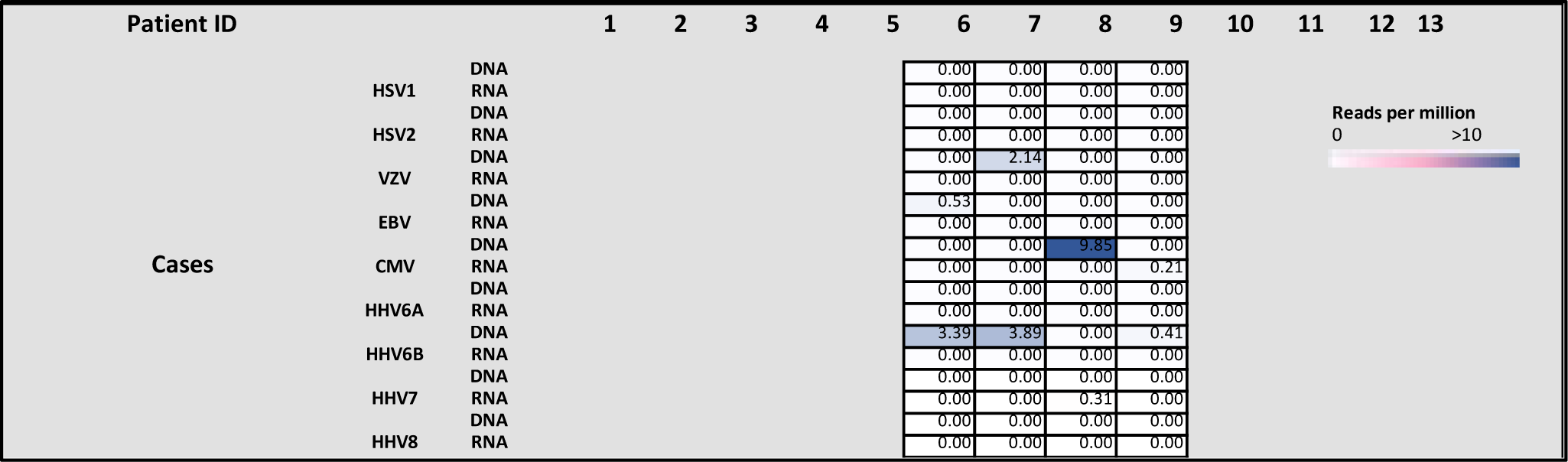
Read counts per million of human herpesviruses in cases following target enrichment sequencing.

**Supplementary Table 5.**
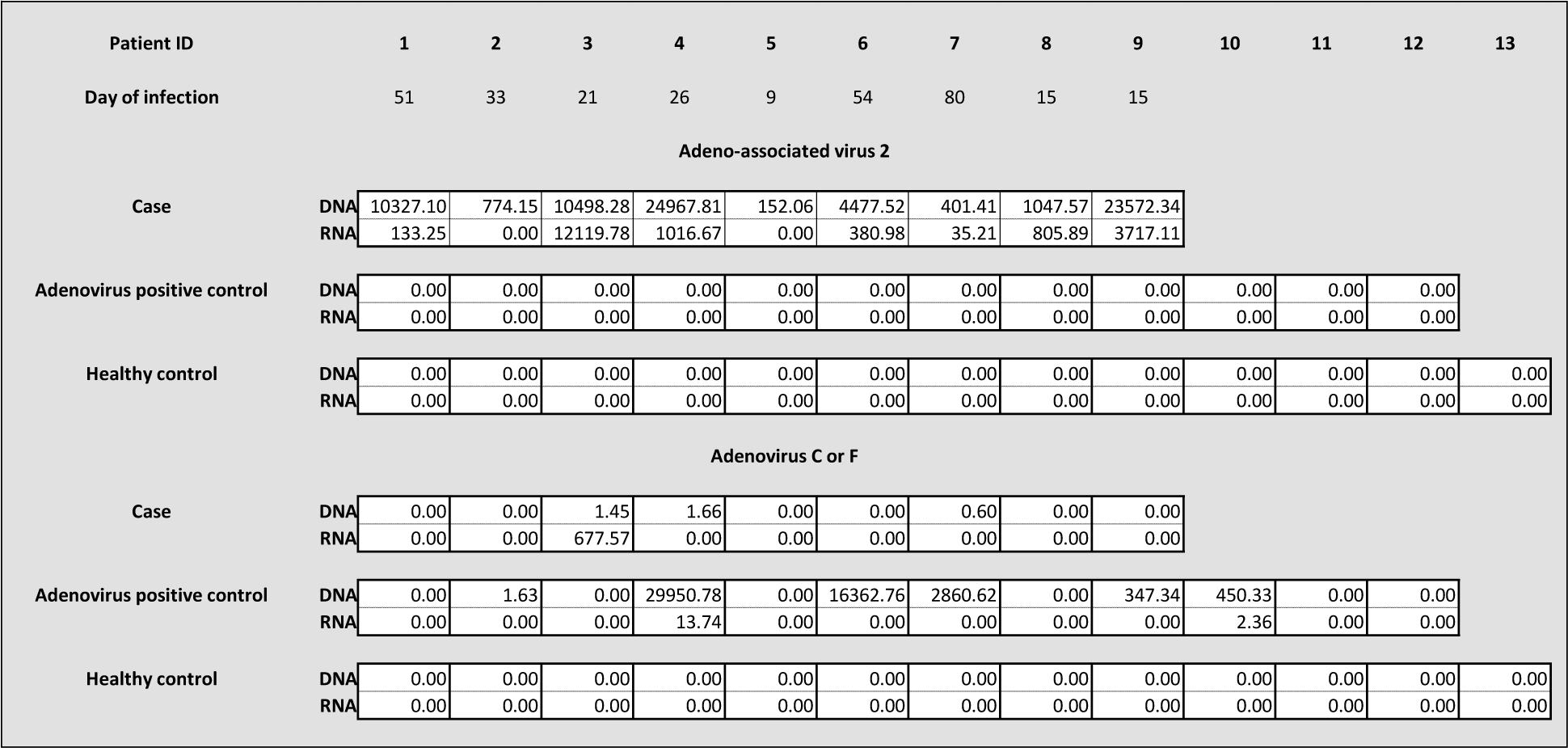
Read counts per million of AAV2 and HAdV in case and control sera following target enrichment sequencing.

**Supplementary Table 6.**
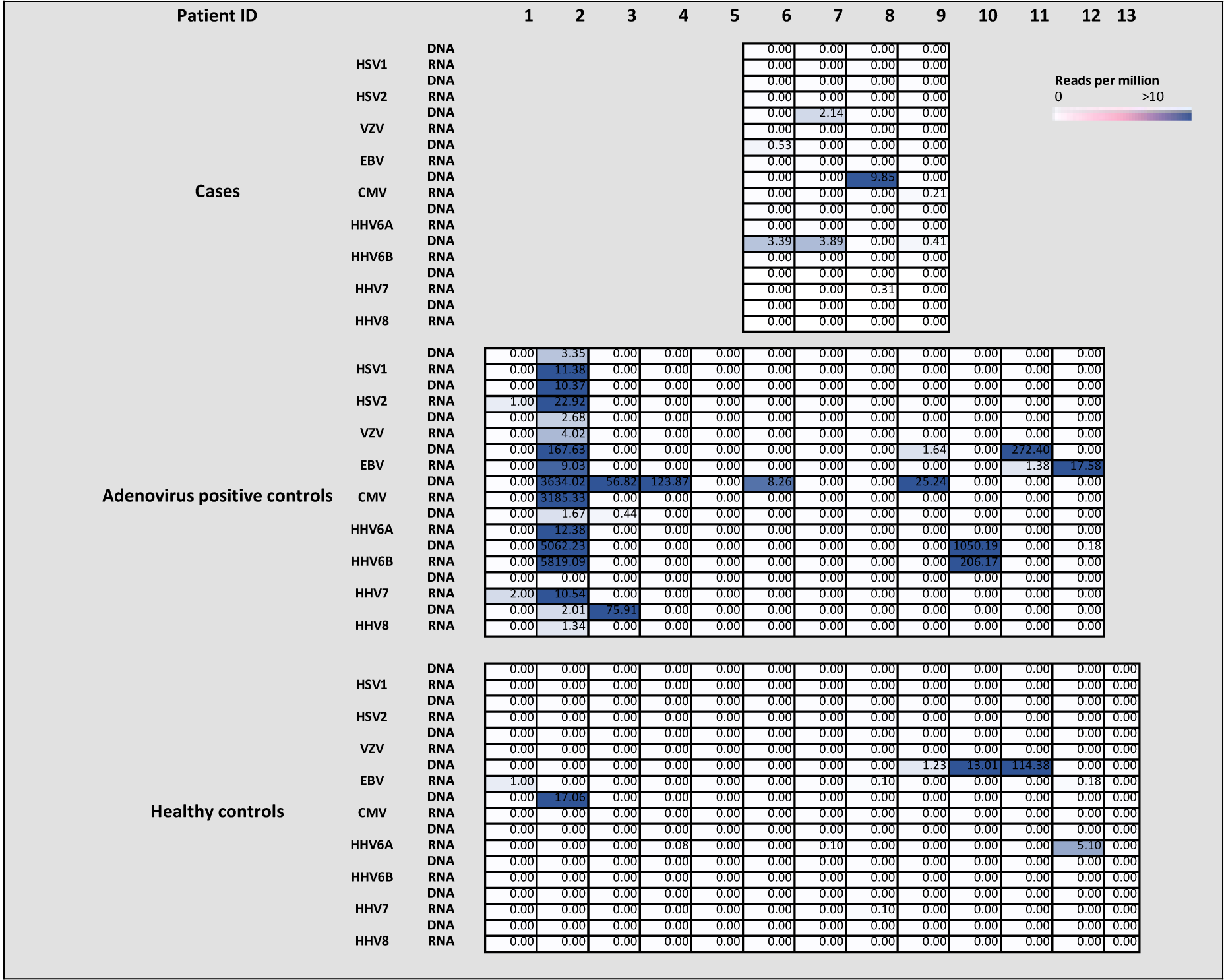
Read counts per million of HHV in case and control sera following target enrichment sequencing.

**Supplementary Figure 1:**
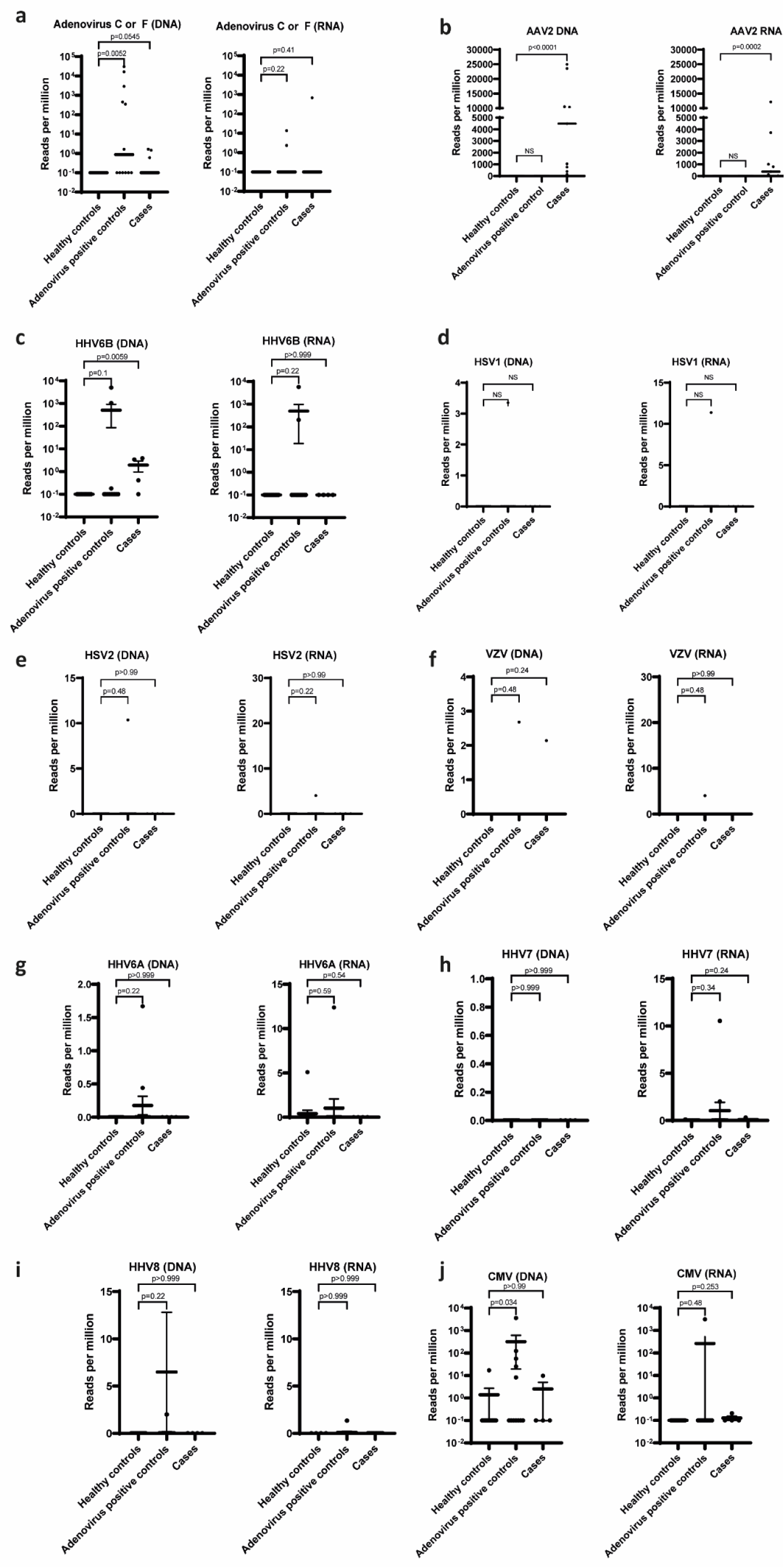
AAV2, HAdV and human herpesvirus detection by target enrichment sequencing in cases and controls. Read counts per million are plotted for HAdV (**a**), AAV2 (**b**), HHV6B (**c**), HSV1 (**d**), HSV2 (**e**), VZV (**f**), HHV6A (**g**), HHV7 (**h**), HHV8 (**i**) and CMV (**j**) in cases, healthy controls and HAdV positive children with normal =

**Supplementary Figure 2:**
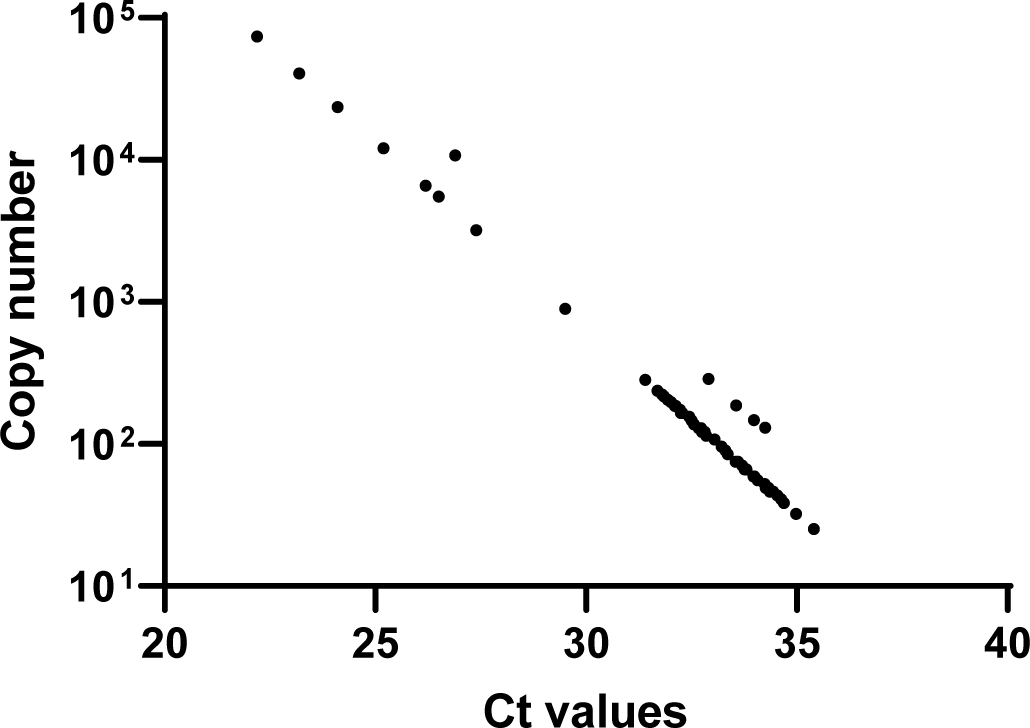
Ct values and estimated copy number for AAV2 real-time. PCR. Estimated copy numbers are shown, calculated using serial dilutions of a plasmid containing the 62bp ITR product to generate a standard curve which was then used to calculate the copy number of AAV2 in samples

**Supplementary Figure 3.**
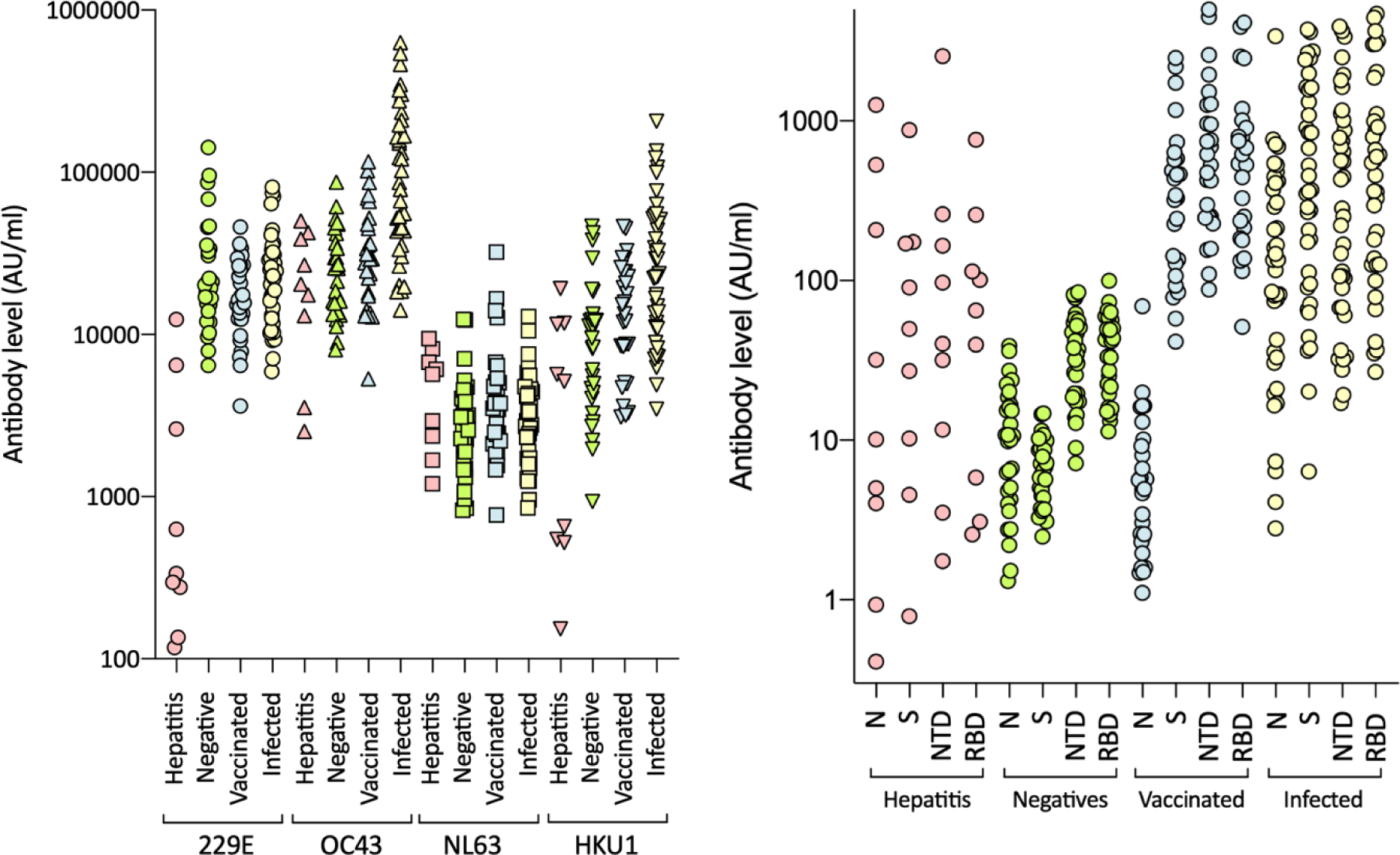
Reactivity to a) human seasonal coronaviruses OC229E, OC43, NL63, HKU1 and b) SARS-CoV-2 in sera from paediatric hepatitis cases. Sera from the paediatric hepatitis cases were screened for reactivity against SARS-CoV-2 nucleocapsid (N), spike (S), N-terminal domain of S (NTD) and receptor binding domain of S (RBD) by electrochemiluminescence (MSD-ECL). Reactivity of the nine samples were compared with three groups of sera from adults of known SARS-CoV-2 status; negatives (n=30), vaccinated two doses (n=28) and infected (n=39).

